# SARS-CoV-2 serology across scales: a framework for unbiased seroprevalence estimation incorporating antibody kinetics and epidemic recency

**DOI:** 10.1101/2021.09.09.21263139

**Authors:** Saki Takahashi, Michael J. Peluso, Jill Hakim, Keirstinne Turcios, Owen Janson, Isobel Routledge, Michael P. Busch, Rebecca Hoh, Viva Tai, J. Daniel Kelly, Jeffrey N. Martin, Steven G. Deeks, Timothy J. Henrich, Bryan Greenhouse, Isabel Rodríguez-Barraquer

## Abstract

Serosurveys are a key resource for measuring SARS-CoV-2 cumulative incidence. A growing body of evidence suggests that asymptomatic and mild infections (together making up over 95% of all infections) are associated with lower antibody titers than severe infections. Antibody levels also peak a few weeks after infection and decay gradually. We developed a statistical approach to produce adjusted estimates of seroprevalence from raw serosurvey results that account for these sources of spectrum bias. We incorporate data on antibody responses on multiple assays from a post-infection longitudinal cohort, along with epidemic time series to account for the timing of a serosurvey relative to how recently individuals may have been infected. We applied this method to produce adjusted seroprevalence estimates from five large-scale SARS-CoV-2 serosurveys across different settings and study designs. We identify substantial differences between reported and adjusted estimates of over two-fold in the results of some surveys, and provide a tool for practitioners to generate adjusted estimates with pre-set or custom parameter values. While unprecedented efforts have been launched to generate SARS-CoV-2 seroprevalence estimates over this past year, interpretation of results from these studies requires properly accounting for both population-level epidemiologic context and individual-level immune dynamics.

## INTRODUCTION

Over the past year, numerous SARS-CoV-2 seroprevalence studies have been conducted to measure population exposure to this novel pathogen (1,2). The need to consider basic assay performance characteristics (sensitivity and specificity) to accurately interpret serosurvey results has been well-established (3–5). Accurate estimation of seroprevalence relies on adequate characterization of assay sensitivity to detect prior infections in the general population. However, for most commercially available assays, manufacturer-reported performance characteristics are usually only applicable to early convalescent samples from hospitalized patients; notably, antibody responses in these individuals are not representative of antibody responses in the general population.

Sufficiently accounting for SARS-CoV-2 antibody responses varying as a function of disease severity (6,7) and waning over time (8,9) is necessary to correctly interpret data from serosurveys performed using these assays.

We previously raised this issue and performed simulations to demonstrate how relying on validation samples that do not represent the distribution of severity and time since infection in a population can introduce spectrum bias into seroprevalence estimation (10). Various modeling approaches have since been proposed to reduce the effects of spectrum bias stemming from antibody waning over time and seroreversion on various serologic platforms (11–15). A key advance of our approach is the ability to parametrize seroreversion using longitudinal antibody kinetic data generated from the same assays used in large-scale serosurveys. To our knowledge, differential antibody responses by disease severity (and factors associated with disease severity such as age (16)) have not yet been incorporated alongside these temporal considerations into a unified framework to accurately estimate seroprevalence from surveys. Failing to account for factors that reduce assay sensitivity will typically underestimate the cumulative SARS-CoV-2 attack rate in the population (10).

Here, we present a flexible statistical approach to produce adjusted seroprevalence estimates that incorporate assay-specific test performance characteristics by severity and time (**Figure 1**). To inform parametrization of the magnitude and kinetics of SARS-CoV-2 immune responses, we used data from a post-infection cohort study with some of the commercial serologic platforms that have been most widely used throughout the pandemic (17). We apply this approach to re-analyze large-scale serosurveys from five locales: Italy, Spain, the United States, Manaus, Brazil, and Japan. Broadly, incorporating variability in individual-level immune dynamics into population-level epidemiologic estimates allows for more accurate estimation of the attack rate, which opens the way for more accurate characterization of population exposure, transmission dynamics, and infection fatality ratios.

**Figure 1:**
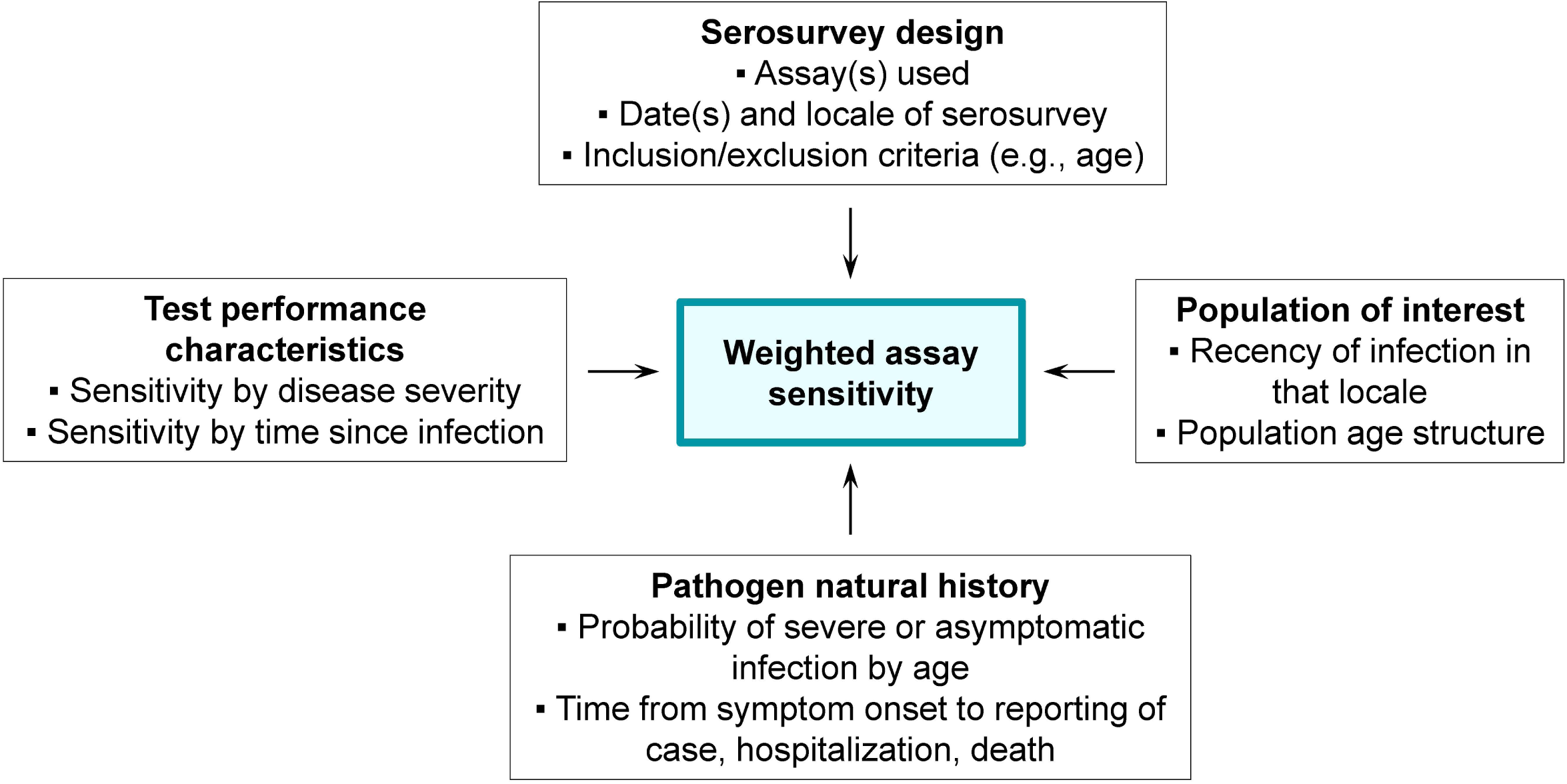
Schematic of the seroprevalence estimation framework. Each of the four boxes on the perimeter details its contributions to the target output of weighted assay sensitivity (center).

## METHODS

### Estimating time-varying, severity-specific assay sensitivities

To estimate time-varying, severity-specific assay sensitivities, we used longitudinal antibody response data collected from a cohort of participants with PCR-confirmed SARS-CoV-2 through the University of California, San Francisco-based *Long term Impact of Infection with Novel Coronavirus* (LIINC) natural history study (NCT04357821). Extensive descriptions of the cohort and laboratory results, including antibody responses on 14 commercial and research-use assays, are available elsewhere (17–19). Briefly, we re-analyzed data published in (17) to estimate assay sensitivity as a function of disease severity and time since symptom onset (Supplementary Table 1). As in (17), we calibrated the time-metric from days since symptom onset (or days since positive PCR test, for asymptomatic individuals) to days since expected seroconversion by adding 21 days to the former (20). For parsimony, we equated having had severe disease with having required hospitalization (16,21).

Building off of the approach we described in (17), individuals were partitioned into 2 severity groups depending on whether or not they were hospitalized for their SARS-CoV-2 infection. We modeled log-transformed signal to cutoff (S/C) or cutoff index (COI) values using Bayesian linear mixed effects models (**Figure 2**; see Supplementary Methods) (22). We estimated sensitivity by severity group and assay continuously for up to 1 year following expected seroconversion (Supplementary Table 2). In sensitivity analyses, we further partitioned non-hospitalized individuals into separate severity groups by asymptomatic and symptomatic. We also extended these methods to estimate joint assay sensitivity from data where every sample was tested on two assays for which results may be correlated (see Supplementary Methods).

**Figure 2:**
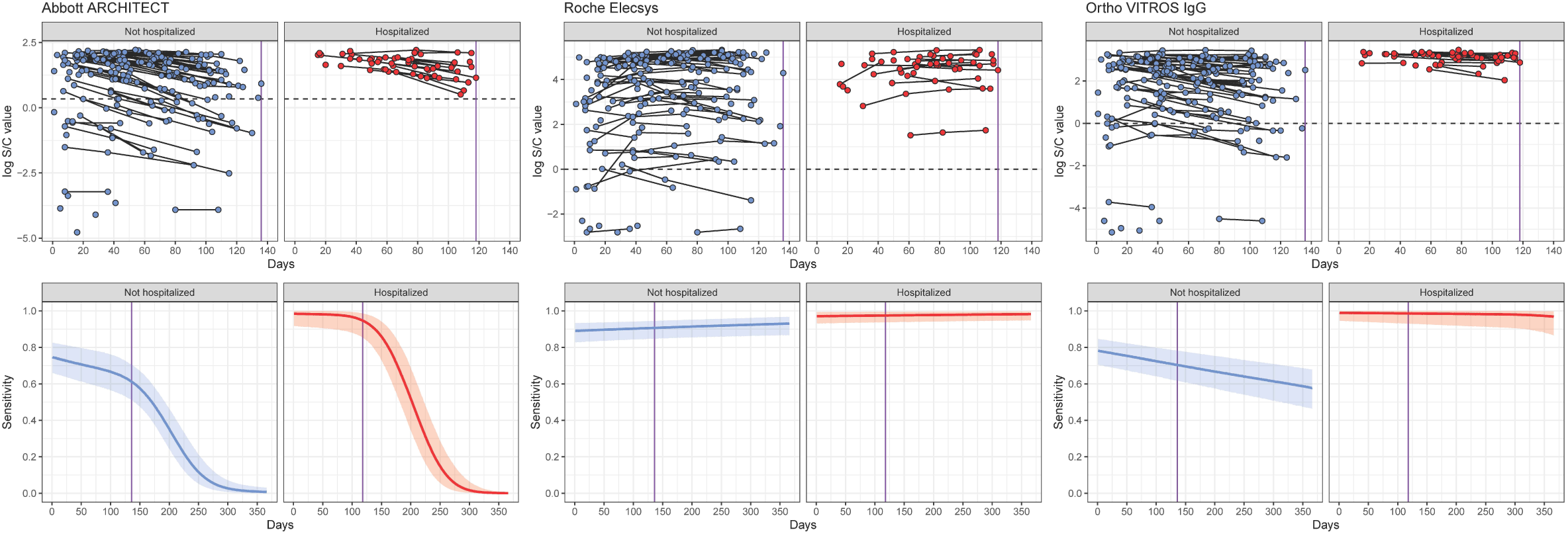
Longitudinal SARS-CoV-2 antibody kinetics and estimated assay sensitivities by time and hospitalization status. (*Upper row*) Time since symptom onset (offset by 21 days) is shown on the x-axis versus the log-transformed antibody response for each of the Abbott ARCHITECT, Roche Elecsys, and Ortho VITROS IgG assays, stratified by hospitalization status. For asymptomatic individuals, the time since the first positive PCR test (offset by 21 days) was used. This time-metric is referred to as ‘time since seroconversion’ hereafter. Longitudinal samples are connected by black lines. Black dotted lines indicate cutoff values for positivity on that assay. (*Lower row*) Estimated sensitivity of each assay (showing posterior median estimates as the solid line and 95% credible intervals), stratified by hospitalization status, from 0 to 365 days after seroconversion.The dashed vertical line in purple indicates the maximum observed time on the corresponding panel above (i.e., x=136 for non-hospitalized and x=118 for hospitalized).

### SARS-CoV-2 serosurveys for application of adjustment framework

The published large-scale serosurveys re-analyzed here were conducted in five locales (23–30). As outlined in **Table 1**, multiple differences exist between these serosurveys in terms of study design, timing, spatial scale, and testing strategies. We included large-scale serosurveys performed using assays with demonstrable heterogeneities in antibody responses, as they would be most affected by issues of spectrum bias (10).

**Table 1:**
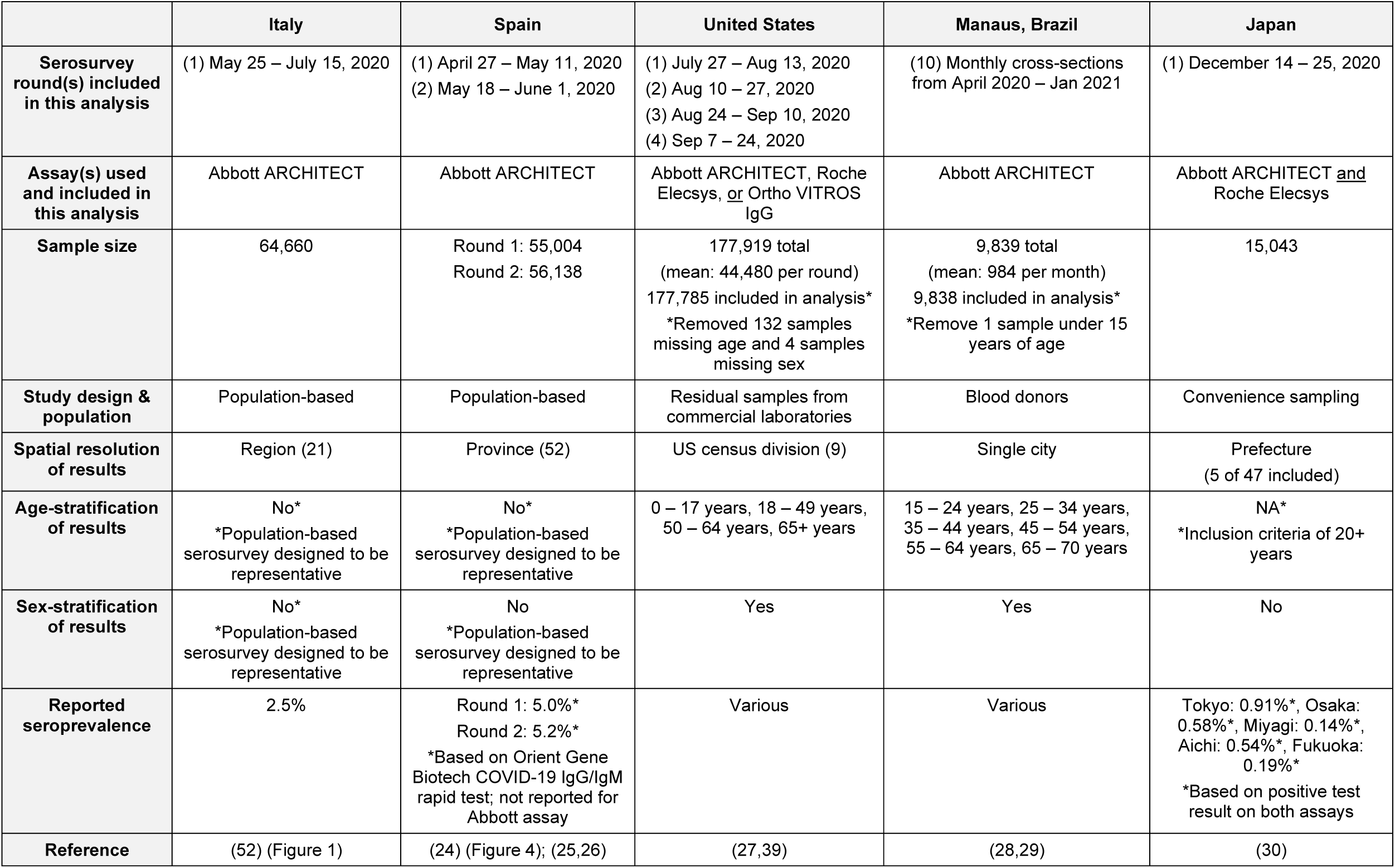
SARS-CoV-2 population serosurveys included.

### Reconstructing time series of symptom onset dates in the locations of selected serosurveys

Properly accounting for the effects of time-varying assay sensitivity on seroprevalence estimates requires understanding how recently individuals in that population might have been infected, relative to when the serosurvey was done. Various data sources can be used as a proxy for population exposure, including time series counts of symptom onsets, positive tests, hospitalizations, and deaths (31). Here, we obtained publicly available time series for each serosurvey locale (**Table 2**) and used symptom onsets as the time-metric for both assay sensitivity and epidemic time series. Where only reporting dates -- but not date of symptom onset -- was available (i.e., United States, Manaus, and Japan), we applied a back-calculation procedure to reconstruct time series of daily symptom onsets using the EpiNow2 software package (32) and parameter estimates for the relevant time delay distributions (33–35) (see Supplementary Methods; Supplementary Table 3). Back-calculation was not necessary for the Italy and Spain time series, as they directly report case counts by date of symptom onset. We also shifted the (reconstructed) time series of symptom onsets forward by 21 days to account for the time between symptom onset and seroconversion.

**Table 2:** SARS-CoV-2 epidemic time series data sets included.

|  | Italy | Spain | United States | Manaus, Brazil | Japan |
| --- | --- | --- | --- | --- | --- |
| **Data type (frequency)** | Reported symptom onsets (daily)*<br>*Region-level data are of positive tests and are available from February 24, 2020 onwards | Reported symptom onsets (daily) | Reported cases and deaths (daily) | Reported hospitalizations (daily) | Reported cases and deaths (daily) |
| **Time delay distribution(s) needed to obtain symptom onset time series** | NA | NA | Case/death report to symptom onset | Hospitalization report to symptom onset | Case/death report to symptom onset |
| **Date of first available data point** | January 28, 2020 | January 1, 2020 | January 21, 2020 | March 16, 2020 | January 16, 2020 |
| **Spatial resolution** | Region | Province | State | Single city | Prefecture |
| **Reference** | Istituto Superiore di Sanità, EpiCentro (53,54) | Gobierno de España, Centro Nacional de Epidemiología (55) | New York Times (56) | Fundação de Vigilância em Saúde do Amazonas - FVS/AM (57,58) | National-level data sets from the Ministry of Health, Labour and Welfare (59,60)<br>Prefectural-level data sets from prefectural ministries of health (61–65) |

Sub-national time series in Italy, Spain, and Japan were generally congruent to national time series during waves of infection included here, so we used a single national-level time series to represent recency of infection for these serosurveys (Supplementary Figures 1-3). Assay-specific seroprevalence data in the United States were only available at the census division level, so we aggregated state-level time series to the census division using weights derived from the number of samples tested by state and by survey round (Supplementary Figure 4). As available, we compared results of symptom onset reconstruction from multiple sources of data (Supplementary Figures 5-6).

### Joint framework for obtaining adjusted seroprevalence estimates

For each serosurvey, we first calculated a single time-varying assay sensitivity, obtained as the average of the severity-specific, time-varying sensitivities, weighted by the expected distribution of disease severities among the serosurvey population. We considered the age distribution of participants in the serosurvey and combined this with published estimates on age-specific weights for the expected proportions of asymptomatic, non-hospitalized, and hospitalized infections (Supplementary Tables 4-6, Supplementary Figure 7). Although a degree of variation in population age structure exists across the included serosurveys, the resulting distribution of severities was, on average, 5% hospitalized (severe), 45% symptomatic and not hospitalized, and 50% asymptomatic.

To obtain an estimate of the expected sensitivity of the assay at the time of a serosurvey, we calculated the dot product of the severity-weighted time-varying assay sensitivity and the difference between the reconstructed time series of symptom onsets and the date of the serosurvey. Using the posterior distribution of this single weighted sensitivity that accounts for both severity and time, along with the manufacturer reported point estimates of specificity for each assay (Supplementary Table 2), we obtained adjusted seroprevalence estimates and 95% credible intervals using the Rogan-Gladen estimator (3) or a binomial model of seroprevalence (4). A multinomial model (36,37) was used for the two-assay scenario. We used the R statistical software (version 3.5.3), EpiNow2 R package (version 1.2.1), and the Stan programming language (versions 2.19.3 and 2.21.2) for all analyses.

## RESULTS

### Kinetics of antibody responses and time-varying, severity-specific assay sensitivity

Across each of the three assays included here (Abbott ARCHITECT, Roche Elecsys, and Ortho VITROS IgG), average antibody responses in hospitalized individuals were consistently higher than those in non-hospitalized individuals, and thus sensitivity estimates were higher in this group (**Figure 2**). Antibody responses in non-hospitalized individuals were strikingly heterogeneous: some individuals had high responses on par with hospitalized individuals, while others had distinctly lower responses. Antibody responses and estimated sensitivities on the Abbott ARCHITECT assay and, to a lesser extent, the Ortho VITROS IgG assay decayed over time, while they remained stable on the Roche Elecsys assay. Additional time points tested suggest that these trends persist over subsequent months (Supplementary Figure 8). Though limited by a small sample size in our study, assay sensitivity for asymptomatic individuals may be substantially lower than that for non-hospitalized, symptomatic individuals (Supplementary Figure 9). We identified similar antibody waning rates on the Abbott ARCHITECT assay in longitudinal samples from the blood donor population in Manaus used for SARS-CoV-2 serosurveillance (Supplementary Figure 10).

Since the serosurveys in Japan performed parallel testing on the Abbott ARCHITECT and Roche Elecsys assays, we jointly modeled the probability of testing positive on both assays. We found that this was highest in the earliest times since infection; the probability of testing negative on Abbott and positive on Roche increased over time, consistent with relatively rapid declines in sensitivity over time on the Abbott assay and consistently high sensitivity of the Roche assay (Supplementary Figure 11). Parameter values for all fitted antibody kinetics models are provided in Supplementary Tables 7-9.

### Overall impacts of serosurvey timing and demography on expected assay sensitivity

Timings of serosurveys relative to the local epidemic curve varied. In some settings, serosurveys were conducted a few months after the first peak, while others during or after periods of ongoing transmission. **Figure 3** shows reconstructed daily numbers of symptom onsets relative to the timing of each serosurvey locale. These are based on the reported time series identified in Table 2 and delay distributions identified in Supplementary Table 3.

**Figure 3:**
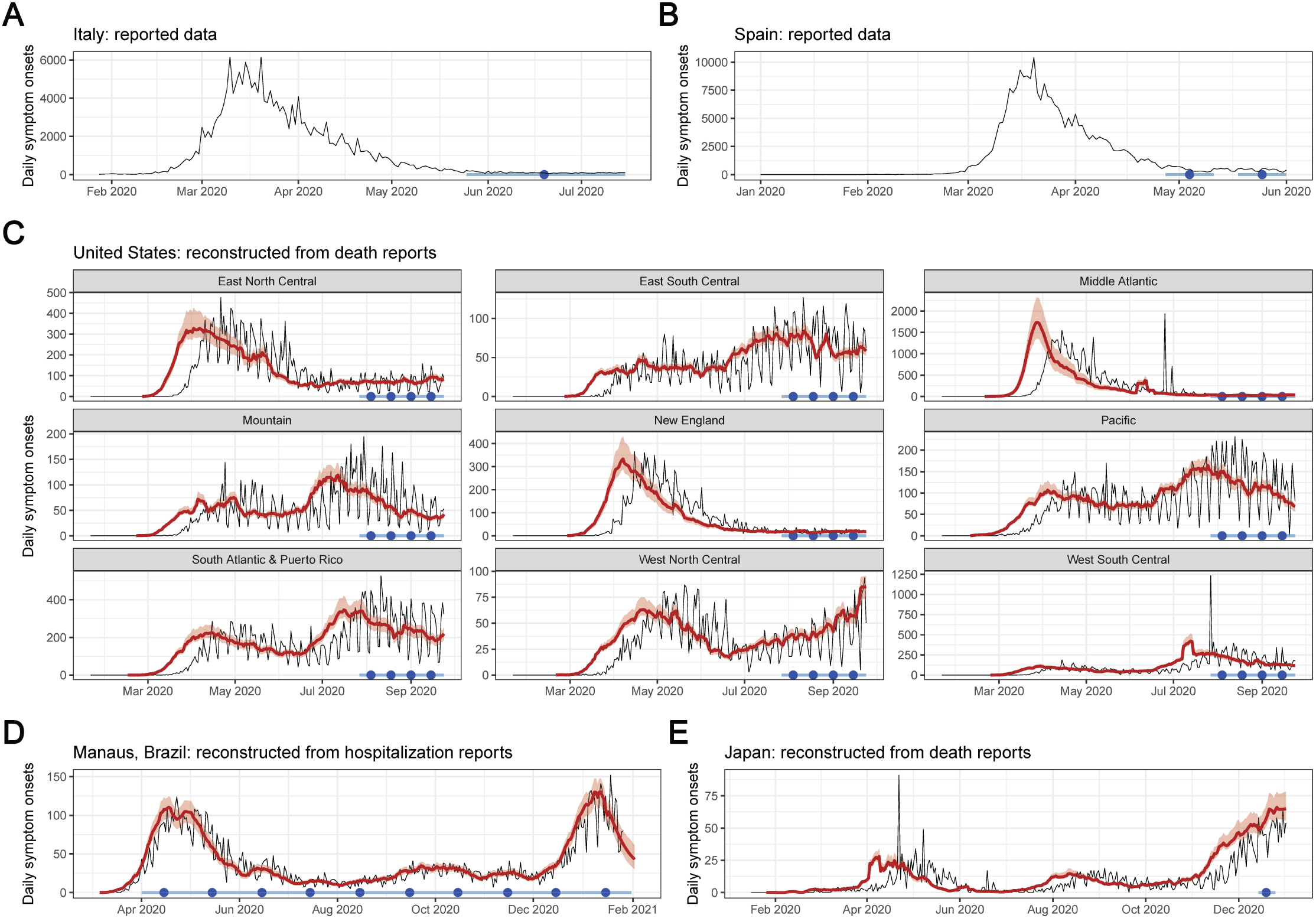
Reported epidemic time series from serosurvey locales, reconstructed symptom onsets, and serosurvey timing. Serosurvey dates are shown by the light blue bar (range) and dark blue circle (midpoint). For reconstructed symptom onset curves, reported data are shown as the black curve, and posterior median estimates and 95% credible intervals are shown in red. **(A)** Reported daily symptom onsets in Italy, January to July 2020. **(B)** Reported daily symptom onsets in Spain, January to June 2020. **(C)** Daily symptom onsets by census division in the United States, reconstructed from death reports, February to September 2020. **(D)** Daily symptom onsets in Manaus, Brazil, reconstructed from hospitalization reports, March 2020 to January 2021. **(E)** Daily symptom onsets in Japan, reconstructed from death reports, January to December 2020. Reconstructed symptom onsets from death reports precede deaths by approximately 3 weeks, while reconstructed symptom onsets from hospitalization reports precede hospitalizations by approximately 10 days.

Estimates of expected assay sensitivity that account for local epidemic recency, antibody waning, and disease severity differed considerably from manufacturer reported sensitivity values (**Table 3**). Expected sensitivity was lower when surveys were performed in locations where infections occurred longer ago, particularly for serosurveys using the Abbott assay. The serosurvey in Italy, the serosurvey rounds in Spain, and the June 2020 round of the serosurvey in Manaus were all conducted at similarly recent times relative to their local epidemics, resulting in similar expected sensitivities of around 70% based on the 2 severity group model. In contrast, serosurveys conducted longer ago relative to their local epidemic had lower expected sensitivities (e.g., the estimate was as low as 45% for the December 2020 round of the serosurvey in Manaus). Expected sensitivity was also strongly influenced by our choice of model, and in particular whether asymptomatic individuals were included with other non-hospitalized individuals or classified in a separate group (Supplementary Figure 12).

**Table 3:**
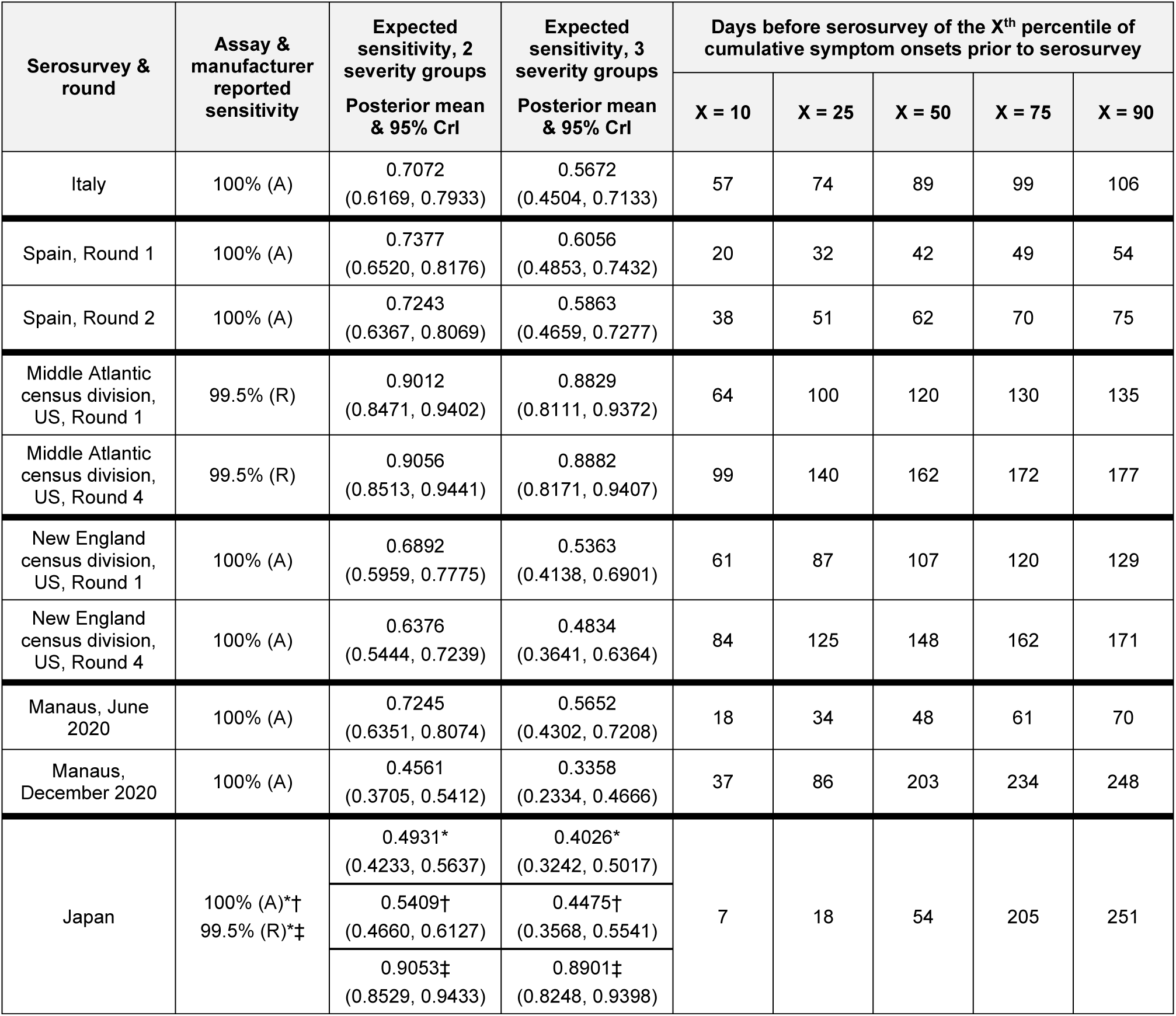
Comparison of manufacturer reported assay sensitivity and expected sensitivity for selected rounds of each serosurvey, and the timing of the serosurvey relative to the local epidemic. The rightmost 5 columns represent percentiles, in days before serosurvey. For example, in Italy, 10% of all symptom onsets occurring prior to the serosurvey occurred within the 57 days before the serosurvey, and 90% of all symptom onsets occurring prior to the serosurvey occurred within the 106 days before the serosurvey. A = Abbott ARCHITECT; R = Roche Elecsys. ^*^Sensitivity is defined as the probability of testing positive on both assays, given infection. †Sensitivity is defined as the probability of testing positive on the Abbott assay, given infection (from a univariate model). ‡Sensitivity is defined as the probability of testing positive on the Roche assay, given infection (from a univariate model).

### Adjusted seroprevalence estimates and degree of potential spectrum bias by survey

#### Italy

The raw national seroprevalence result of this population-based serosurvey, conducted between May 25 and July 15, 2020, was 2.5% using the Abbott ARCHITECT assay. Based on our 2 severity group model, we estimated adjusted seroprevalence to be 3.0% (95% credible interval (CrI): 2.7%, 3.4%).Based on our 3 severity group model, adjusted seroprevalence increased to 3.7% (95% CrI: 3.0%, 4.7%). As identified in the original serosurvey report, northern regions of the country were particularly affected during the first wave of infection (**Figure 4A**). As we set the weighted assay sensitivities to be equal across regions, the ratio of adjusted to raw seroprevalence scaled with raw seroprevalence: in the Lombardia region (orange on Figure 4A), which had the highest raw seropositive proportion at 7.5% in this survey, the adjusted seroprevalence was estimated to be 1.35-fold greater (95% CrI: 1.20, 1.55) than reported (**Figure 5A**; Supplementary Figure 13). Ratios below 1 represent regions with extremely low raw seroprevalence, accounting for expected test performance (i.e., false positive results due to imperfect specificity, see Supplementary Methods).

**Figure 4:**
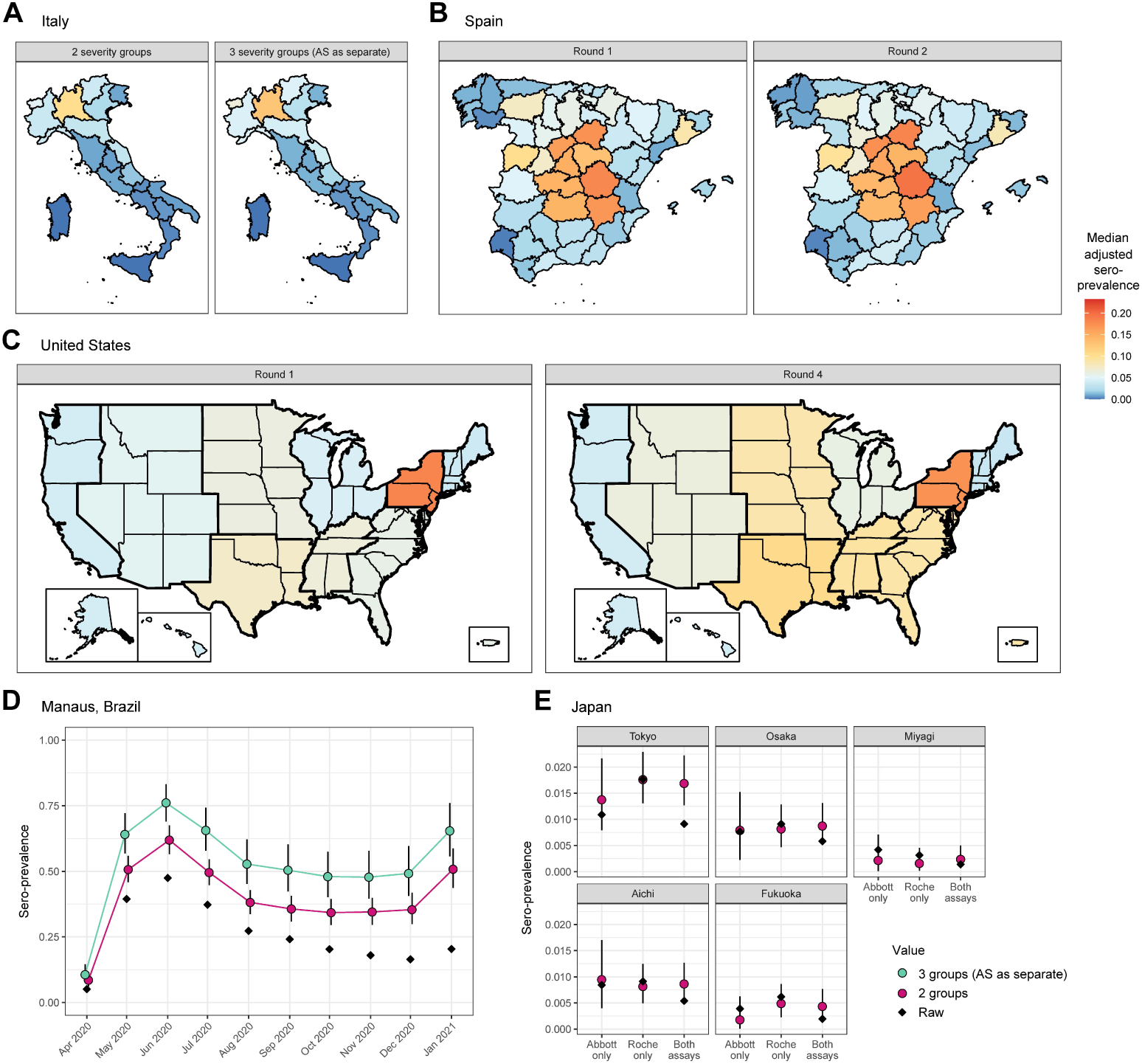
Adjusted seroprevalence estimates by survey. Panels A, B, and C all share the same color scale. **(A)** Estimated seroprevalence by region in Italy, using the antibody kinetics model classifying disease severity into (*left*) 2 groups: non-hospitalized and hospitalized, and (*right*) 3 groups: asymptomatic (AS), symptomatic and non-hospitalized, and hospitalized. The former is considered as the primary scenario for this analysis, and the subsequent panels are derived under that scenario unless otherwise described. **(B)** Estimated seroprevalence by province in Spain for (*left*) Round 1 and (*right*) Round 2 of the serosurvey. **(C)** Estimated seroprevalence by census division in the United States for (*left*) Round 1 and (*right*) Round 4 of the serosurvey, incorporating weighting by the population demographics (age and sex) of each census division, as well as age-specific probabilities of hospitalization. **(D)** Estimated seroprevalence by month in Manaus, Brazil, incorporating weighting by age and sex, as well as age-specific probabilities of hospitalization and of experiencing symptoms, restricted to between the ages of 15 and 70 years. The raw seropositive proportion is in black; results from the antibody kinetics model classifying disease severity into 2 groups are in pink, and 3 groups are in green. **(E)** Estimated seroprevalence for the 5 prefectures in Japan, considering the raw results from the (*left*) Abbott assay only, (*center*) Roche assay only, and (*right*) both assays. The raw seropositive proportion is in black and the estimated seroprevalence is in pink. When considering the results from both assays, the raw seropositive proportion is the proportion of samples that tested positive on both.

**Figure 5:**
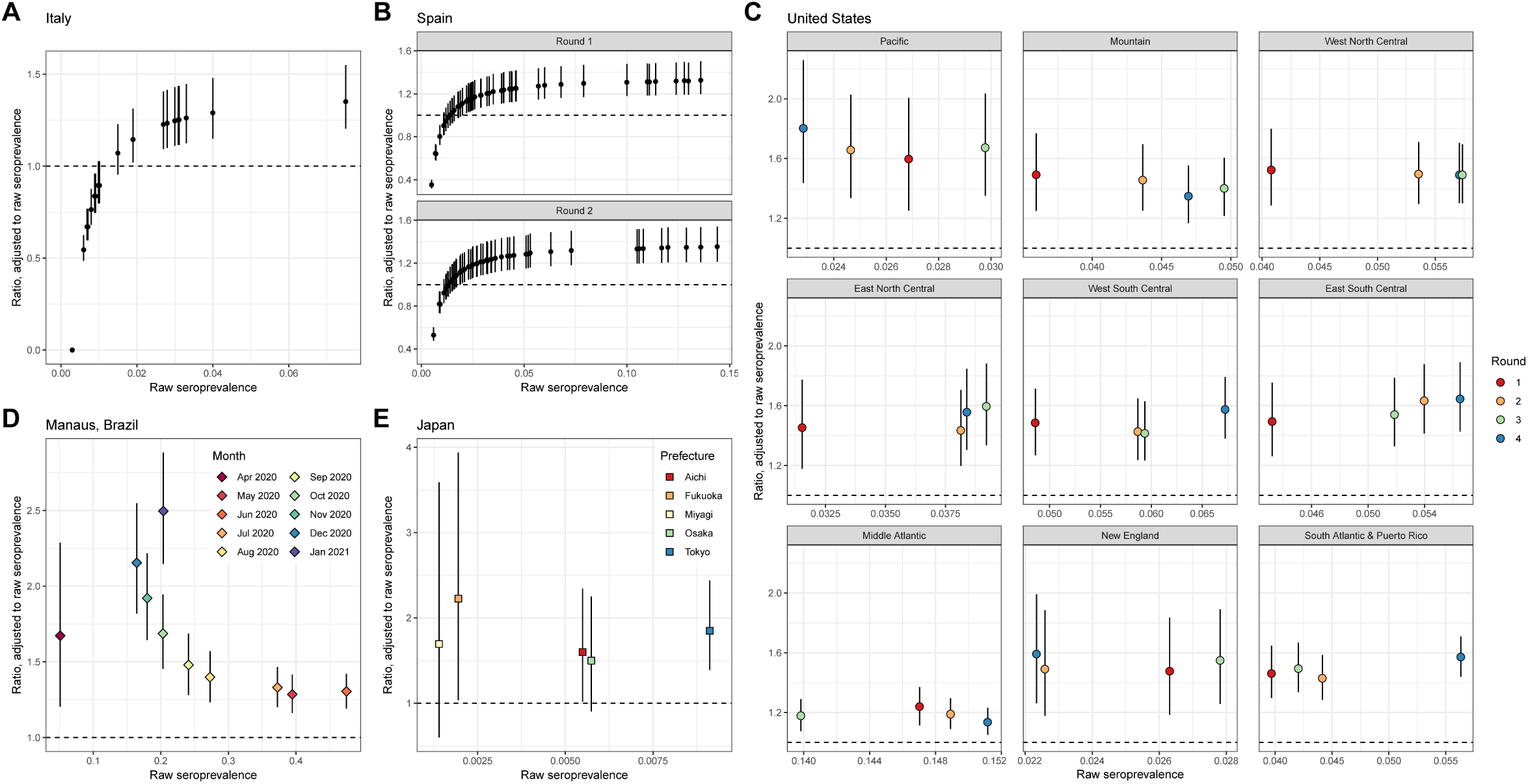
Relative bias in seroprevalence estimation. For each panel, the raw seroprevalence result is shown on the x-axis and the ratio of the adjusted to raw seroprevalence is shown on the y-axis (median and 95% credible interval). The ratio equaling 1 (i.e., no bias) is shown in the dashed line. All panels are generated under the primary scenario of classifying disease severity into 2 groups, non-hospitalized and hospitalized. **(A)** Italy, where each point represents a region. **(B)** Spain, for (*upper*) Round 1 and (*lower*) Round 2, where each point represents a province. **(C)** The 9 census divisions of the United States, where the color of the point represents the survey round. **(D)** Manaus, Brazil, where each point represents a month. As in Figure 4, for panels C and D, the adjusted seroprevalence estimates are weighted by population demography and age-specific disease severity. **(E)** Japan, where each point represents a prefecture. The scenario considered here is the case of using the results of the two assays.

#### Spain

The raw national seroprevalence results for this population-based serosurvey, conducted in two rounds, were 5.0% (Round 1: April 27-May 11, 2020) and 5.2% (Round 2: May 18-June 1, 2020) using a rapid serologic test, and 4.6% (Round 1) and 4.5% (Round 2) using the Abbott ARCHITECT assay. Point estimates of seroprevalence were generally consistent within provinces between rounds (**Figure 4B**; Supplementary Figure 14), which may be explained by their close temporal spacing (early May and late May 2020). Based on our 2 severity group model with the Abbott ARCHITECT data, we estimated adjusted seroprevalence to be 5.8% (95% CrI: 5.2%, 6.5%) in Round 1 and 5.7% (95% CrI: 5.2%, 6.5%) in Round 2. The ratio of adjusted to raw seroprevalence within a province and serosurvey was similar to in Italy, reaching a high of 1.35-fold increase (95% CrI: 1.21, 1.54) over the reported value in the Cuenca province (dark orange on Figure 4B) (**Figure 5B**). For additional context, the adjusted seroprevalence estimates in Round 2 here, using the 3 severity group model, were as high as what was measured (using a rapid test) during Round 4 of the survey (7.1%), which was conducted from November 16 to 29, 2020 (38).

#### United States

The first four rounds of the US CDC Nationwide Commercial Laboratory Seroprevalence Survey were conducted between July and September 2020, and were performed on multiple serologic assay platforms at the census division level (Supplementary Figures 15-18) (27,39). Adjusted point estimates at the census division level from Round 1 and Round 4 using our 2 severity group model are provided in **Figure 4C**, and estimates from the other rounds are available in Supplementary Figure 19. Compared to the raw seroprevalence results, adjusted seroprevalence estimates were up to 2-fold greater (**Figure 5C**; Supplementary Figures 20-21). Aggregation of data from multiple serologic assay platforms complicates interpretation of the raw results. For instance, in the Mountain census division, which comprises 8 states, all three assays were used; without further information, it is not possible to know which states used which combinations of the three. On the other hand, all of the data from the Middle Atlantic states (New York, New Jersey, and Pennsylvania) were from the Roche Elecsys assay (which exhibited the most stable antibody responses over time of the three assays), and thus the biases in this census division are the lowest in the United States (Figure 5C).

To further explore the relative effects of time and assay choice on weighted sensitivity, we simulated serosurveys at various times at the census division level using different proportions of tests conducted on the Abbott ARCHITECT assay (i.e., decreasing responses over time), assuming that the rest of the tests were done on the Roche Elecsys assay (i.e., stable responses over time) (**Figure 6**). In general, we found that a greater proportion of tests performed on the Abbott assay leads to a lower weighted sensitivity. However, even if a serosurvey was performed exclusively with one assay and serosurveys were conducted at the same time in all census divisions, expected assay sensitivity in the general population will differ considerably between census divisions due to the differential timings of the epidemic in the population. For example, a serosurvey conducted February 2021 in the Middle Atlantic census division using exclusively the Abbott assay would have an expected sensitivity of 32%, while a serosurvey in the West North Central census division at the same time using the same assay would have an expected sensitivity of 59%.

**Figure 6:**
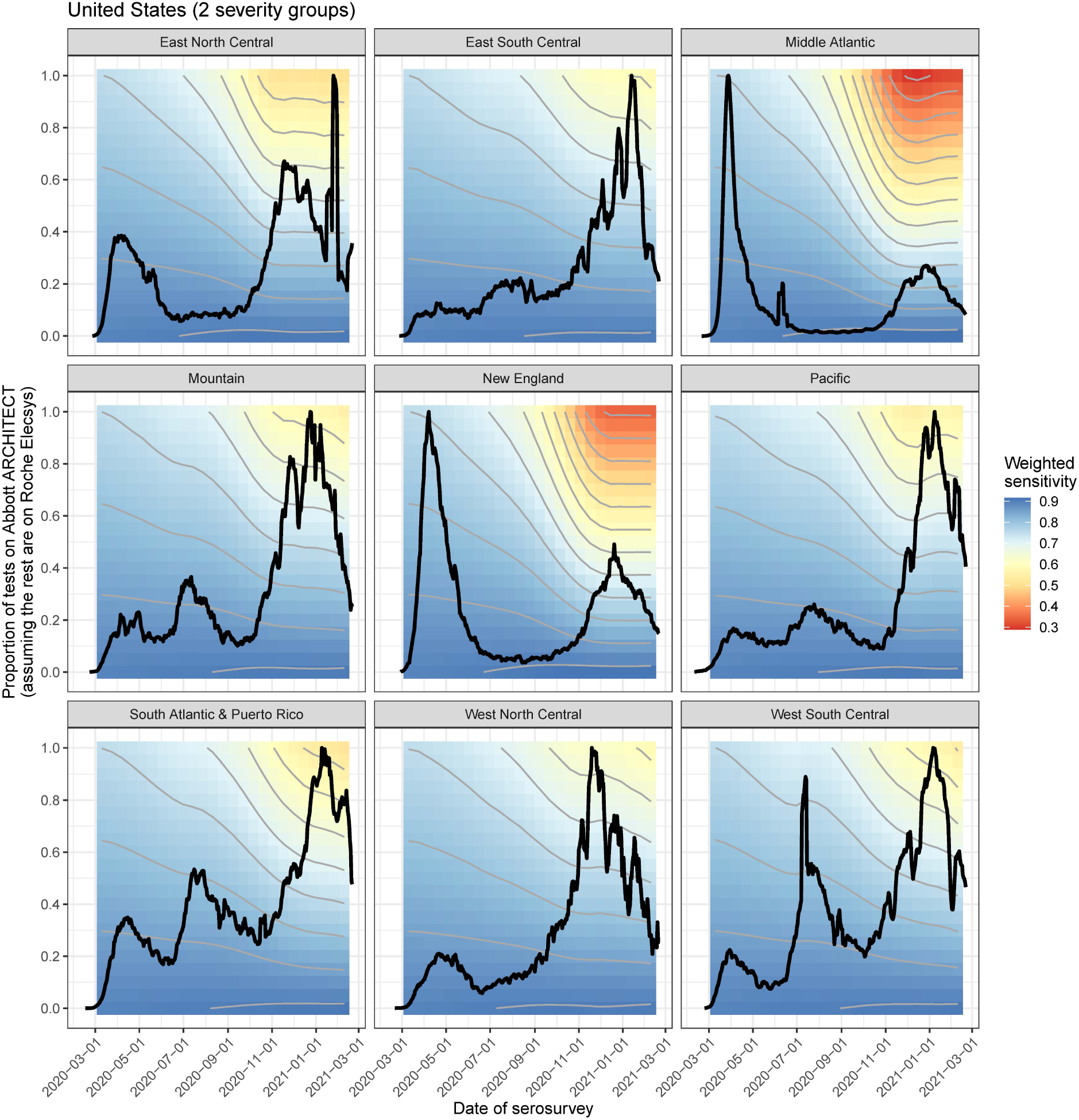
Weighted sensitivity by time and assays used for the US CDC Nationwide Commercial Laboratory Seroprevalence Survey. Weighted assay sensitivity by census division in the United States as a function of (hypothetical) serosurvey dates between March 2020 and March 2021 (x-axis), and proportion of tests performed on the Abbott ARCHITECT assay (y-axis), incorporating symptom onset curves (black line) and demography at the census division level. This is using the 2 severity group scenario. This assumes that the rest of the tests were performed on the Roche Elecys assay, on which antibody responses remain robust over time. Contour interval of 0.05.

#### Manaus, Brazil

Seroprevalence in Manaus, Brazil has been measured monthly since April 2020 in cross-sectional samples from blood donors using the Abbott ARCHITECT assay. Manaus experienced a particularly large first wave of the SARS-CoV-2 pandemic in Spring 2020, followed by a relative trough period (Figure 3D). Both raw and adjusted seroprevalence peaked in June 2020 (raw seroprevalence of 47.5% and estimated seroprevalence of 61.9% (95% CrI: 56.5%, 67.5%) using the 2 severity model) and then declined thereafter (**Figure 4D**). Temporal changes in the ratio of adjusted to raw seroprevalence echo the epidemic dynamics in Manaus, where the degree of bias increased over time from 1.28-fold (95% CrI: 1.16, 1.42) in May 2020 to 2.50-fold (95% CrI: 2.15, 2.88) in January 2021 (**Figure 5D**; Supplementary Figure 22).

#### Japan

In the convenience sampling-based serosurveys in 5 prefectures in Japan conducted in December 2020, each sample was tested on both the Abbott ARCHITECT and the Roche Elecsys platforms (30). The raw seroprevalence values varied considerably between the univariate (single assay) and bivariate interpretations (in the latter, a sample had to be positive on both assays to be deemed a positive). We found adjusted seroprevalence values to be generally comparable between the univariate and bivariate models, but using either only Roche Elecsys or both assays led to lower uncertainty than using the Abbott ARCHITECT assay alone (**Figure 4E**). Based on our bivariate model with 2 severity groups, the ratio of adjusted to raw seroprevalence ranged from 1.50-fold (95% CrI: 0.90, 2.25) in Osaka prefecture to 2.23-fold (95% CrI: 1.04, 3.94) in Fukuoka prefecture (**Figure 5E**; Supplementary Figure 23). Importantly, having all samples in this serosurvey tested on two assays, along with all validation sets also being tested on both assays, provided the opportunity to estimate the correlation between the assays (36) and to incorporate this into the estimation (see Supplementary Methods).

## DISCUSSION

Differences in antibody responses as a function of disease severity and time since infection complicate inference from population-level serologic data, because they can substantially affect the expected sensitivity of the assay. In the serosurveys evaluated here, we found that true seroprevalence was up to 2.5-fold greater than that measured by raw seropositivity. Leveraging data on the kinetics of antibody responses measured by three of the most widely used serologic assays, we present a unified methodological framework to estimate SARS-CoV-2 seroprevalence that accounts for these factors and provide a toolkit for practitioners to generate adjusted seroprevalence estimates with either pre-set or custom parameter values.

We found that the degree of bias depends on the assay used, the timing of the serosurvey relative to the course of the epidemic locally, and the age distribution of the population and the age-dependent probability of severe disease. The serosurvey in Italy and the two serosurvey rounds in Spain were conducted relatively early with respect to the local epidemic, resulting in expected sensitivities of around 70%. In the United States, the serosurveys included here occurred during the first and second waves of the pandemic and used different assays in different census divisions, affecting the magnitude of the bias in estimated seroprevalence. In the Middle Atlantic census division, the magnitude of the bias was attenuated due to the exclusive use of the Roche assay; in the New England census division, which used the Abbott assay exclusively, the magnitude of the bias was greater. In Japan, the serosurvey was conducted almost a year after the first case of COVID-19 was reported (40). While the absolute magnitudes of both raw and adjusted seroprevalence were low (under 2%), expected sensitivity on the Abbott assay for this serosurvey was as low as 54%.

In Manaus, the magnitude of the bias continuously increased from 1.28-fold in May 2020 to 2.50-fold in January 2021, revealing the footprint of the first wave of the pandemic in early 2020. These results corroborate findings that seroprevalence was likely already high prior to the subsequent resurgence of SARS-CoV-2 infection in late 2020 (15,29). The decrease in adjusted seroprevalence estimates after June 2020 suggest that these adjustments for severity and time may be insufficient, as we do not expect actual cumulative incidence to decrease over time. This could be attributable to issues of sampling, age-patterns not adequately captured in this framework, or higher waning rates. The framework developed here provides an alternative to a previously developed approach (15). Our framework does not impose assumptions on seroprevalence estimates monotonically increasing at each month, but rather assumes that reported case counts (in this instance, hospitalizations) are an accurate reflection of temporal trends in transmission. Our framework implicitly assumes temporal homogeneity in the case fatality ratio, case hospitalization ratio, and the case reporting ratio. Known deviations from this assumption through estimation of time-varying metrics could be incorporated to improve the accuracy of estimates (e.g., potential decreases in the case fatality ratio over time (41)).

This approach is broadly applicable beyond the assays and serosurveys included. We previously presented raw data to estimate time-varying sensitivity of 11 additional assays using cohort data (17), and numerous studies have also studied longitudinal antibody responses on an array of platforms (42–44). A key methodological development here is in providing a framework to use these data to adjust for time since infection as informed by the local epidemic in a population of interest. This framework incorporates a number of setting-specific scenarios, including the availability of different types of reported epidemiologic data, population representativeness of the serosurvey and necessary adjustments for weights, spatial scale, study design, and testing strategy.

Two of the assays included here, the Abbott ARCHITECT and Ortho VITROS IgG, have considerable waning over the first 5 months following infection. Persistence of these trends will lead to further decreases in assay sensitivity. Over time, antibody waning will be increasingly important to account for, as more individuals will have been infected longer ago and with greater variability. These considerations will be important for interpreting subsequent rounds of serosurveys included here (e.g., Spain (38) and the United States (39)) and others not included (e.g., India (45)). These considerations underscore the need for continued longitudinal follow-up of individuals with confirmed SARS-CoV-2 infection (and various strains), as antibody kinetics often follow more complex dynamics of boosting and waning over time beyond linear changes (46). Assays demonstrating more waning may be better suited for other use-cases such as identifying recent infections. An additional consideration for designing a serosurvey in places where mRNA vaccines are used is on using assays measuring antibodies to non-spike proteins, which will play a role in distinguishing immune responses to natural infection from vaccine-elicited immune responses to the spike protein alone.

There are a number of caveats associated with this analysis. The accuracy of this approach hinges on the accuracy of symptom onset curves reconstructed from the selected reported time series. This limitation is not unique to seroprevalence estimation; accurate estimates of downstream metrics such as the time-varying reproduction number similarly rely on the robustness of these data streams over time (32,47). Estimation will also be sensitive to the data type chosen; for example, hospitalizations and deaths are generally more robust to temporal trends in under-ascertainment than cases, but this may be context-specific.

A key consideration here is the small sample size for asymptomatically infected individuals, who potentially comprise a majority of all SARS-CoV-2 infections (48,49). While the distinction between asymptomatic versus minimally symptomatic may be difficult to define, it is imperative to better understand the magnitude and kinetics of antibody responses in this group of individuals to better understand the extent of bias in seroprevalence estimates. The decision to model asymptomatic individuals as their own severity group or aggregated with the other non-hospitalized individuals has a major effect on overall adjusted seropositivity. Our framework accounts for differences in assay sensitivity by disease severity and time, but does not explicitly incorporate other potentially important sources of variation, such as age and sex (16,50,51). Lastly, our focus has been on sensitivity, and we do not allow for specificity to vary over time.

This work provides a broadly applicable framework incorporating individual-level immune dynamics into epidemiologic models to produce adjusted seroprevalence estimates for a number of serosurvey-specific scenarios. The methodology has been made publicly available for broad public use (https://github.com/sakitakahashi/spectrum-bias-adjust). More accurate seroprevalence estimates will allow for better understanding of the proportion that has been exposed to date, and for various applications including integration into downstream mechanistic transmission models.

## Data Availability

Raw antibody data used in this analysis are publicly available in a prior publication (17). Raw serosurvey data are available in the citations in Table 1. Access to COVID-19 seroprevalence data from the Nationwide Commercial Laboratory Seroprevalence Survey is maintained by the Centers for Disease Control and Prevention's (CDC)'s Epi Task Force Seroprevalence Team. Requests for access to the data should be directed to. The CDC does not take responsibility for the scientific validity or accuracy of methodology, results, statistical analyses, or conclusions presented.

## ACKNOWLEDGEMENTS

ST is supported by the Schmidt Science Fellows, in partnership with the Rhodes Trust. ST, IR, and IRB acknowledge research funding from the MIDAS Coordination Center COVID-19 Urgent Grant Program (MIDASNI2020-5), by a grant from the National Institute of General Medical Science (3U24GM132013-02S2). IRB is supported by R35GM138361-02. We acknowledge Dr. Kristina Bajema for sharing the assay-specific data from the first 4 rounds of the CDC serosurvey.

## DATA AND CODE AVAILABILITY

Raw antibody data used in this analysis are publicly available in a prior publication (17). Raw serosurvey data are available in the citations in Table 1. Access to COVID-19 seroprevalence data from the Nationwide Commercial Laboratory Seroprevalence Survey is maintained by the Centers for Disease Control and Prevention’s (CDC)’s Epi Task Force Seroprevalence Team. Requests for access to the data should be directed to. The CDC does not take responsibility for the scientific validity or accuracy of methodology, results, statistical analyses, or conclusions presented. All code to reproduce these analyses are available at: https://github.com/sakitakahashi/spectrum-bias-adjust.

