## Supplementary Information for "SARS-CoV-2 serology across scales: a framework for unbiased seroprevalence estimation incorporating antibody kinetics and epidemic recency"

**Table of Contents**

Supplementary Methods

Supplementary Tables 1-9

Supplementary Figures 1-23

Supplementary References

### Supplementary Methods

#### Modeling SARS-CoV-2 antibody kinetics and assay sensitivity: single assay (univariate)

For observation $h$ for individual $i$ of disease severity group $s\in\{1,2\}$ (primary scenario of 2 disease severity groups: non-hospitalized and hospitalized) or $s\in\{1,2,3\}$ (secondary scenario of 3 disease severity groups: asymptomatic, symptomatic and non-hospitalized, and hospitalized), we modeled their log-transformed S/C antibody response $Y_{h,i}$ on each assay in a Normal Bayesian hierarchical model as follows:

$${p(Y}_{h,i}|u_{i},\lambda,Time_{h,i},\sigma)=Normal(u_{i}+\lambda\cdot Time_{h,i},\sigma)$$

$${p(u}_{i}|s,\theta,\beta,\tau)=\theta_{s}\cdot Normal(u_{i}|\beta_{1}, \tau_{1}) + {(1-\theta}_{s})\cdot Normal(u_{i}|\beta_{2}, \tau_{2})$$

Above,$u_{i}$ represents an individual-level random effect arising from a mixture of two distributions representing low and high responders. $\theta_{s}$ represents the probability of being a low responder in disease severity group $s$. The low responder mixture component is parametrized as a Normal distribution with mean $\beta_{1}$ and standard deviation $\tau_{1}$, and the high responder mixture component is parametrized as a Normal distribution with mean $\beta_{2}$ and standard deviation $\tau_{2}$. The decision to model the individual-level random effects using mixture distributions was driven by the observations of antibody responses in non-hospitalized individuals being highly heterogeneous, with some individuals in this group having high responses on par with hospitalized individuals, and others having distinctly lower responses; we modeled the random effects in hospitalized individuals using mixture mixtures for reciprocity. In estimation we constrained $\tau_{1}\geq\tau_{2}$ to ensure non-exchangeability between mixtures.

In addition, $\lambda$ represents the fixed effect of $Time_{h,i}$, where $Time_{h,i}$ is data on the time since symptom onset (if symptomatic) or since positive PCR test (if asymptomatic). For parsimony, we assumed that $\lambda$ (which is interpreted as the antibody decay or growth rate) did not change over time, and assumed that this rate is shared across all individuals. $e_{h,i}$ represents the residual error that is normally distributed with a mean of 0 and a standard deviation of $\sigma$.

To estimate changes in assay sensitivity over time, we simulated population distributions of $\hat{Y}$ for disease group $s$ by iteratively sampling values from each of the posterior distributions of the parameters $\lambda,\theta_{s},\beta,\tau,\sigma$. We assumed the linear trends to hold for up to 1 year (i.e., simulated values in $0\leq Time\leq365$), which is consistent with longitudinal serologic data from additional studies [[1]](https://paperpile.com/c/CEaFMo/Jj48a). For each sampled value of the parameters, we then sampled draws of $u_{i}$ and $e_{hi}$ and determined the assay sensitivity for disease severity group $s$ at time $t$ days after seroconversion ($Se_{s,t}$) as the proportion of overall draws where $\hat{Y}$ was above the log-transformed assay cutoff value for positivity. We used the law of total probability to calculate time-varying sensitivity for an assay, weighted by disease severity ($Se_{t}$), where disease severity $p(s)$ was estimated in Supplementary Tables 5 and 6:

$$Se_{t}=\sum_{s=1}^{S} Se_{s,t}\cdot p(s)$$

We ran 4 Markov chain Monte Carlo (MCMC) chains of length 50,000 each using the Stan programming language (<https://mc-stan.org/>), and assessed convergence using the Gelman-Rubin $\hat{R}$ statistic. We used uninformative priors for all parameters and hyper-parameters.

#### Modeling SARS-CoV-2 antibody kinetics and assay sensitivity: two assays (bivariate)

We extended the univariate models for antibody kinetics and assay sensitivity above (with two disease severity groups $s\in\{1,2\}$) to incorporate the scenario where each sample was tested on two assays, on which results may be correlated. We modeled the paired log-transformed S/C antibody response $Z_{h,i}$ on each assay using a Multivariate (bivariate) Normal Bayesian hierarchical model, where $Z_{h,i}$ is a two-dimensional vector representing results on assays $a\in\{1,2\}$ as follows:

${p(Z}_{h,i}|\mu,\Sigma)=MultiNormal(\mu,\Sigma)$ ⇒ $\mu_{a}=u_{a,i}+\lambda_{a}\cdot Time_{h,i}$

$${p(u}_{a,i}|s,\theta,\beta,\tau)=\theta_{a,s}\cdot Normal(u_{a,i}|\beta_{a,1}, \tau_{a,1}) + {(1-\theta}_{a,s})\cdot Normal(u_{a,i}|\beta_{a,2}, \tau_{a,2})$$

Above, $\Sigma$ is a $2 \times2$ covariance matrix, assumed to be fixed over time. In estimation we constrained $\tau_{a,1}\geq\tau_{a,2}$ to ensure non-exchangeability between mixtures. We estimated the following three severity-specific, time-varying bivariate assay sensitivities using the simulation approach described for the univariate scenario, but now for $\hat{Z}$:

$$Se_{1,s,t}=p(Abbott+, Roche+|s,t)$$

$$Se_{2,s,t}=p(Abbott+, Roche-|s,t)$$

$$Se_{3,s,t}=p(Abbott-, Roche+|s,t)$$

$$Se_{4,s,t}=p(Abbott-, Roche-|s,t)=1-(Se_{1,s,t}+Se_{2,s,t}+Se_{3,s,t})$$

We again used the law of total probability to calculate time-varying sensitivity for an assay, weighted by disease severity ($Se_{1,t}$, $Se_{2,t}$, $Se_{3,t}$, and $Se_{4,t}$):

$$Se_{1,t}=\sum_{s=1}^{S} Se_{1,s,t}\cdot p(s)$$

$$Se_{2,t}=\sum_{s=1}^{S} Se_{2,s,t}\cdot p(s)$$

$$Se_{3,t}=\sum_{s=1}^{S} Se_{3,s,t}\cdot p(s)$$

$$Se_{4,t}=\sum_{s=1}^{S} Se_{4,s,t}\cdot p(s)$$

#### Reconstructing symptom onset time series from reported data

Based on the reported time series $D_{t}$ (e.g., daily counts of death reports) and the time delay distribution $f(\tau)$ (e.g., probability of $\tau$days between symptom onset to death reporting, Supplementary Table 3), we used the EpiNow2 software to reconstruct the time series of symptom onsets, $C_{t}$, which are unobserved:

$${p(D}_{t}|C_{t},f(\tau),\omega,\psi)=NegBin\left( \sum_{\tau=0}^{t} C(t-\tau)\cdot f(\tau)\cdot\omega_{t mod 7},\psi) \right)$$

Above, $\omega_{t mod 7}$ represents a categorical ‘day of the week’ effect with an independent parameter for each day of the week to account for potential heterogeneities in reporting, and $\psi$ represents the overdispersion parameter of the Negative Binomial observation model [[2]](https://paperpile.com/c/CEaFMo/gruD). Note that when the reported time series are of symptom onsets, $D_{t}=C_{t}$ and reconstruction is not necessary. Bootstrapped log-Normal distributions of $f(\tau)$ were generated using EpiNow2.

#### Estimating an overall weighted assay sensitivity

Two pieces of information are used to estimate an overall weighted assay sensitivity $Se$ that is ultimately used to adjust a serosurvey result:

1) Estimated time-varying sensitivity (weighted by severity in the population of interest) $Se_{t}$, where $0\leq t\leq365$ represents days since symptom onset. $S{e_{t}}^{*}=Se_{t+21}$ represents the shifted sensitivity curve, incorporating an additional 21 days between symptom onset and expected seroconversion.
2) Time series of symptom onsets $C_{t}$, where $0 \leq t \leq T$. Here, $t=0$ represents the first date where $C>0$, and $T$ represents the number of days between the corresponding date of $t=0$ and the midpoint date of the serosurvey, minus 21 days in order to account for individuals who had been infected by the time of the serosurvey but not yet seroconverted. Lastly, the scaled value ${c_{t}}^{*}=\frac{C_{t}}{\sum C_{t}}$ is used to ensure the weighted assay sensitivity is between 0 and 1.

$S{e_{t}}^{*}$ is truncated to $({{Se}_{0}}^{*},{Se_{1}}^{*},...,{{Se}_{T}}^{*})$ so that the time-varying sensitivity and the symptom onset curve ${c_{t}}^{*}$are the same length. The final overall weighted assay sensitivity, $Se_{wtd}$, is estimated as the dot product of $S{e_{t}}^{*}$ and the reverse of ${c_{t}}^{*}$, i.e., $reverse({c_{t}}^{*})=({c_{T}}^{*},{c_{T-1}}^{*},...,{c_{0}}^{*}).$ The exact same procedure is used in the bivariate scenario to estimate $Se_{wtd,1}$, $Se_{wtd,2}$, $Se_{wtd,3}$, and $Se_{wtd,4}$.

#### Estimating adjusted SARS-CoV-2 seroprevalence

Using the posterior of weighted assay sensitivity that accounts for both severity and time ($Se_{wtd}$), as well as the reported point estimate of assay specificity ($Sp$), we obtained adjusted seroprevalence estimates and 95% credible intervals using the following methods:

1) In a univariate scenario: if raw seropositivity was reported as a percentage ($p_{raw}$), then by using the Rogan-Gladen estimator [[3]](https://paperpile.com/c/CEaFMo/OqIf1):

$p_{adj}=\frac{p_{raw} + Sp - 1}{Se_{wtd} + Sp - 1}$

The Rogan-Gladen estimator can also be rearranged to determine when the ratio of adjusted to raw seroprevalence will be less than 1 (e.g., Figure 5 in the main text):

$p_{adj}<p_{raw}$ ⇒ $p_{raw}<\frac{1 - Sp}{1 - (Se_{wtd} + Sp - 1)}$

2) In a univariate scenario: if the numerator ($x$) and denominator ($N$) counts of the raw serosurvey data are available, then the Binomial distribution can be used to estimate the adjusted seroprevalence [[4,5]](https://paperpile.com/c/CEaFMo/wTqE+DXpP). For the US serosurveys, where multiple assays were sometimes used, we assumed the existence of a single overall $p_{adj}$ rather than assay-specific seroprevalence:

$p(x|N,p_{adj})=Binomial(N,p_{adj}\cdot Se_{wtd} +(1-p_{adj})\cdot(1-Sp))$

3) In a bivariate scenario: the Binomial model above can be generalized to the two-assay scenario using a Multinomial model of seroprevalence [[5,6]](https://paperpile.com/c/CEaFMo/8tdUj+DXpP). We can jointly model the raw test results of both assays $x_{ab}$ (e.g., $x_{++}$ represents the number of samples that tested positive on both assays), estimating a single adjusted seroprevalence value $p_{adj}$:

$$p(x_{++},x_{+-},x_{-+},x_{--}|p_{adj,}Se_{wtd,1},Se_{wtd,2},Se_{wtd,3},Se_{wtd_{4}},Sp_{a},Sp_{b})=Multinomial(\alpha)$$

$$\alpha_{1}=p_{adj}\cdot Se_{wtd,1}+(1-p_{adj})\cdot(1-Sp_{a})\cdot(1-Sp_{b})$$

$$\alpha_{2}=p_{adj}\cdot Se_{wtd,2}+(1-p_{adj})\cdot(1-Sp_{a})\cdot Sp_{b}$$

$$\alpha_{3}=p_{adj}\cdot Se_{wtd,3}+(1-p_{adj})\cdot Sp_{a}\cdot(1-Sp_{b})$$

$$\alpha_{4}=p_{adj}\cdot Se_{wtd,4}+(1-p_{adj})\cdot Sp_{a}\cdot Sp_{b}$$

#### Analytical pipeline in the Stan programming language

The three primary steps in the analytical pipeline use the following inputs and outputs:

1) Estimate time-varying, severity-specific assay sensitivities (in Stan)

- **Input:** longitudinal antibody kinetics data
- **Output:** time-varying sensitivity, severity-specific assay sensitivity and posteriors as a Stan object

2) Reconstruct time series of symptom onsets for a given serosurvey (using EpiNow2, in Stan)

- **Input:** raw reported time series data (case, hospitalization, or death reports, primarily)
- **Output:** counts of symptom onsets by date and posteriors as a Stan object

3) Adjusted seroprevalence estimates (in Stan, using generated quantities function gqs())

- **Input:** outputs of Steps 1 and 2, age-specific probabilities of hospitalization and age-specific probabilities of experiencing symptoms, assay specificity
- **Intermediate step 1:** calculate time-varying sensitivity, weighted by severity
- **Intermediate step 2:** normalize counts of symptom onsets by date to sum to 1
- **Intermediate step 3:** obtain a single, overall weighted assay sensitivity by taking the dot product of the severity-weighted, time-varying sensitivity and reverse of the normalized symptom onset time series
- **Output:** adjusted seroprevalence estimate and posteriors

### Supplementary Tables

**Supplementary Table 1: Longitudinal antibody data included from the LIINC cohort study.** Note that we have previously made all of the data referenced here publicly available in [[7]](https://paperpile.com/c/CEaFMo/xEVn).

| **Characteristic** | **N=127** |
| --- | --- |
| **Clinical Manifestations of COVID-19** |  |
| Asymptomatic | 8 |
| Total symptomatic (non-hospitalized + hospitalized) | 119 |
| Symptomatic and hospitalized | 31 |
| **Enrollment and follow-up** |  |
| Follow-up time, days since onset (median) | 112 (range: 22-157) |
| Time points contributed (median) | 2 (range: 1-4) |
| Total samples contributed | 265 |

**Supplementary Table 2: Manufacturer reported test performance characteristics for the three commercial SARS-CoV-2 serologic assays included.** S/C = signal to cutoff index. COI: cutoff index.

|  | **Abbott ARCHITECT SARS-CoV-2 IgG** | **Roche Elecsys Anti-SARS-CoV-2 Total** | **Ortho Clinical Diagnostics VITROS Anti-SARS-CoV-2 IgG** |
| --- | --- | --- | --- |
| **Antigen target** | N protein | N protein | S protein |
| **Sensitivity (at ≥ 2 weeks post-infection)** | 100%  (88/88) | 99.5%  (184/185) | 90%  (36/40) |
| **Specificity (pre-pandemic samples and/or other respiratory illness)** | 99.63%  (1,066/1,070) | 99.80%  (10,432/10,453) | 100%  (407/407) |
| **Cutoff value for positivity** | S/C ≥ 1.4 | COI ≥ 1.0 | S/C ≥ 1.0 |
| **Reference** | [[8]](https://paperpile.com/c/CEaFMo/FOme) | [[9]](https://paperpile.com/c/CEaFMo/tAue) | [[10]](https://paperpile.com/c/CEaFMo/CyvT) |

**Supplementary Table 3: Time delay distributions used for reconstructing time series of symptom onsets.**

|  | **Symptom onset to case report** | **Symptom onset to hospitalization report** | | **Symptom onset to death report** |
| --- | --- | --- | --- | --- |
| **Reference** | [[11]](https://paperpile.com/c/CEaFMo/whVd) (Table S2) | [[12]](https://paperpile.com/c/CEaFMo/nFtz) (Table 2) | [[11]](https://paperpile.com/c/CEaFMo/whVd) (Table S2) | [[12]](https://paperpile.com/c/CEaFMo/nFtz) (Table 2) |
| **Parameter values (days)** | Gamma(2.12, 0.39) | Lognormal  Mean: 9.7  SD: 35.2 | Gamma(1.23, 0.79) | Lognormal  Mean: 20.2  SD: 11.6 |
| **Usage note in this analysis** | Supplementary scenario for US and Japan | Primary scenario for Manaus, Brazil | Supplementary scenario for Manaus, Brazil | Primary scenario for US and Japan |

**Supplementary Table 4: Demographic data sets included.**

|  | **Italy** | **Spain** | **United States** | **Manaus, Brazil** | **Japan** |
| --- | --- | --- | --- | --- | --- |
| **Indicators of interest for this analysis** | Population size by region  Population size by age category (national) | Population size by province  Population size by age category (national) | Population size by census division, age category, sex  Population size by state (for comparison with results in Bajema *et al* [[13]](https://paperpile.com/c/CEaFMo/XoZk)) | Population size by age category, sex | Population size by age category (national) |
| **Reference** | [[14]](https://paperpile.com/c/CEaFMo/2iwd) (Table 1)  Istituto Nazionale di Statistica (2020 data): [[15]](https://paperpile.com/c/CEaFMo/UKDC) | [[16]](https://paperpile.com/c/CEaFMo/CUiP) (Supplementary Table 1) - via Spanish National Institute of Statistics (2019 data) | American Community Survey (2018 data): [[17]](https://paperpile.com/c/CEaFMo/l7gc) | Brazilian Institute of Geography and Statistics (2010 census data): [[18]](https://paperpile.com/c/CEaFMo/6y2w) | Japan National Statistics Center (2016 data): [[19]](https://paperpile.com/c/CEaFMo/rMgC) |

**Supplementary Table 5: Literature search of studies estimating age-specific probabilities of hospitalization and age-specific probabilities of experiencing symptoms, conditional on SARS-CoV-2 infection.**

| **Age group** | **Pr(hosp \| infection)**  Values from [[20]](https://paperpile.com/c/CEaFMo/h3pe) | **Pr(hosp \| infection)**  Values from [[21]](https://paperpile.com/c/CEaFMo/YFAr) | **Pr(hosp)**  Values from [[22]](https://paperpile.com/c/CEaFMo/VczA) | **Pr(symptomatic \| infection)**  Values from [[23]](https://paperpile.com/c/CEaFMo/evbD) |
| --- | --- | --- | --- | --- |
| **0 – 9 years** | 0 | 0.0001 | 0 | 0.29 |
| **10 – 19 years** | 0.000408 |  | 0.0008 | 0.21 |
| **20 – 29 years** | 0.0104 | 0.0005 | 0.0008 | 0.27 |
| **30 – 39 years** | 0.0343 | 0.011 | 0.01 | 0.33 |
| **40 – 49 years** | 0.0425 | 0.014 | 0.019 | 0.40 |
| **50 – 59 years** | 0.0816 | 0.029 | 0.054 | 0.49 |
| **60 – 69 years** | 0.118 | 0.058 | 0.151 | 0.63 |
| **70 – 79 years** | 0.166 | 0.093 | 0.333 | 0.69 |
| **80+ years** | 0.184 | 0.262 | 0.618 |  |

**Supplementary Table 6A: Estimated proportion of infected individuals who would experience severe disease, given local population demography and age-specific probabilities of hospitalization and of experiencing symptoms, excluding the United States.** Estimates for Manaus, where age bins in the reported serosurvey data are narrow but shifted from those in Supplementary Table 5, are obtained by interpolation (Supplementary Figure 7). These values are assumed to be the same between sexes.

|  | **Italy** | **Spain** | **Manaus, Brazil** | **Japan** |
| --- | --- | --- | --- | --- |
| **Probability of severe disease in infected population**  Values from [[21]](https://paperpile.com/c/CEaFMo/YFAr) | 0.04389209 | 0.03821921 | 15 – 24 years: 0.00030  25 – 34 years: 0.00575  35 – 44 years: 0.01250  45 – 54 years: 0.02150  55 – 64 years: 0.04350  65 – 70 years: 0.07550 | 0.05683933*  *Restricting to 20+ years as per serosurvey inclusion criteria |
| **Probability of asymptomatic infection in infected population**  Values from [[23]](https://paperpile.com/c/CEaFMo/evbD) | 0.5553919 | 0.5723913 | 15 – 24 years: 0.760  25 – 34 years: 0.700  35 – 44 years: 0.635  45 – 54 years: 0.555  55 – 64 years: 0.440  65 – 70 years: 0.340 | 0.5050978*  *Restricting to 20+ years as per serosurvey inclusion criteria |

**Supplementary Table 6B: Estimated proportion of infected individuals who would experience severe disease, given local population demography and age-specific probabilities of hospitalization, by age group and census division in the United States.** These values are assumed to be the same between sexes.

| **Probability of severe disease** | **Pacific** | **Mountain** | **West North Central** | **East North Central** | **West South Central** | **East South Central** | **Middle Atlantic** | **New England** | **South Atlantic & Puerto Rico** |
| --- | --- | --- | --- | --- | --- | --- | --- | --- | --- |
| **0 – 17 years** | 0.0001 | 0.0001 | 0.0001 | 0.0001 | 0.0001 | 0.0001 | 0.0001 | 0.0001 | 0.0001 |
| **18 – 49 years** | 0.00825 | 0.00823 | 0.0082 | 0.00824 | 0.00828 | 0.00829 | 0.0083 | 0.00824 | 0.00839 |
| **50 – 64 years** | 0.03833 | 0.03878 | 0.0387 | 0.03866 | 0.03821 | 0.03869 | 0.03844 | 0.03847 | 0.03843 |
| **65+ years** | 0.12202 | 0.11826 | 0.12447 | 0.12224 | 0.1189 | 0.1189 | 0.12555 | 0.12371 | 0.12112 |

**Supplementary Table 6C: Estimated proportion of infected individuals who would experience asymptomatic (AS) infection, given local population demography and age-specific probabilities of experiencing symptoms, by age group and census division in the United States.** These values are assumed to be the same between sexes.

| **Probability of AS infection** | **Pacific** | **Mountain** | **West North Central** | **East North Central** | **West South Central** | **East South Central** | **Middle Atlantic** | **New England** | **South Atlantic & Puerto Rico** |
| --- | --- | --- | --- | --- | --- | --- | --- | --- | --- |
| **0 – 17 years** | 0.75118 | 0.75154 | 0.75099 | 0.75191 | 0.75117 | 0.7518 | 0.7517 | 0.75363 | 0.75195 |
| **18 – 49 years** | 0.66952 | 0.66955 | 0.66994 | 0.66902 | 0.66914 | 0.66841 | 0.66869 | 0.66901 | 0.66772 |
| **50 – 64 years** | 0.46495 | 0.46279 | 0.46316 | 0.46335 | 0.46553 | 0.46324 | 0.4644 | 0.46427 | 0.46445 |
| **65+ years** | 0.32996 | 0.32998 | 0.32956 | 0.32975 | 0.33032 | 0.32981 | 0.3288 | 0.32925 | 0.3292 |

**Supplementary Table 7: Parameter estimates for the 2 severity group assay-specific antibody kinetics models.**

| **Abbott ARCHITECT (2 groups)** | | | |
| --- | --- | --- | --- |
| **parameter** | **posterior mean** | **2.5%** | **97.5%** |
| lambda | 0.009255 | 0.008233 | 0.010319 |
| beta[1] | -0.176814 | -0.962516 | 0.611669 |
| beta[2] | 2.211129 | 2.085707 | 2.332211 |
| tau[1] | 1.826817 | 1.463848 | 2.240172 |
| tau[2] | 0.355784 | 0.26827 | 0.450705 |
| sigma | 0.209985 | 0.186817 | 0.236897 |
| theta[1] | 0.422928 | 0.286159 | 0.581739 |
| theta[2] | 0.03778 | 0.001126 | 0.134388 |
| **Roche Elecsys (2 groups)** | | | |
| **parameter** | **posterior mean** | **2.5%** | **97.5%** |
| lambda | -0.001675 | -0.003484 | 0.000011 |
| beta[1] | 1.893436 | 1.165719 | 2.520631 |
| beta[2] | 4.661161 | 4.424638 | 4.843373 |
| tau[1] | 2.08655 | 1.765635 | 2.48082 |
| tau[2] | 0.234237 | 0.079773 | 0.468235 |
| sigma | 0.359686 | 0.319765 | 0.407068 |
| theta[1] | 0.604088 | 0.468024 | 0.732921 |
| theta[2] | 0.173877 | 0.030855 | 0.371309 |
| **Ortho VITROS IgG (2 groups)** | | | |
| **parameter** | **posterior mean** | **2.5%** | **97.5%** |
| lambda | 0.005765 | 0.004574 | 0.007011 |
| beta[1] | 0.569447 | -0.25683 | 1.28965 |
| beta[2] | 3.246154 | 3.093767 | 3.399025 |
| tau[1] | 2.212649 | 1.84487 | 2.653605 |
| tau[2] | 0.366517 | 0.237903 | 0.499033 |
| sigma | 0.241527 | 0.215573 | 0.271736 |
| theta[1] | 0.558846 | 0.40771 | 0.723032 |
| theta[2] | 0.038632 | 0.001153 | 0.135343 |

**Supplementary Table 8: Parameter estimates for the 3 severity group assay-specific antibody kinetics models.**

| **Abbott ARCHITECT (3 groups)** | | | |
| --- | --- | --- | --- |
| **parameter** | **posterior mean** | **2.5%** | **97.5%** |
| lambda | 0.009291 | 0.008244 | 0.010347 |
| beta[1] | -0.24464 | -1.06055 | 0.513569 |
| beta[2] | 2.204515 | 2.075227 | 2.332377 |
| tau[1] | 1.820997 | 1.46352 | 2.284447 |
| tau[2] | 0.368057 | 0.281231 | 0.467742 |
| sigma | 0.209379 | 0.18624 | 0.235204 |
| theta[1] | 0.793623 | 0.471347 | 0.988785 |
| theta[2] | 0.362538 | 0.229402 | 0.506152 |
| theta[3] | 0.035724 | 0.001112 | 0.128795 |
| **Roche Elecsys (3 groups)** | | | |
| **parameter** | **posterior mean** | **2.5%** | **97.5%** |
| lambda | -0.00171 | -0.00342 | 0.000099 |
| beta[1] | 1.830826 | 1.087693 | 2.500225 |
| beta[2] | 4.636431 | 4.396404 | 4.839333 |
| tau[1] | 2.098162 | 1.756128 | 2.505745 |
| tau[2] | 0.266554 | 0.092001 | 0.50027 |
| sigma | 0.360861 | 0.321419 | 0.405696 |
| theta[1] | 0.748399 | 0.435887 | 0.974176 |
| theta[2] | 0.569869 | 0.418818 | 0.702016 |
| theta[3] | 0.161435 | 0.023448 | 0.359104 |
| **Ortho VITROS IgG (3 groups)** | | | |
| **parameter** | **posterior mean** | **2.5%** | **97.5%** |
| lambda | 0.005762 | 0.004552 | 0.007028 |
| beta[1] | 0.561134 | -0.27109 | 1.28038 |
| beta[2] | 3.245625 | 3.079303 | 3.408247 |
| tau[1] | 2.211076 | 1.857575 | 2.64292 |
| tau[2] | 0.365011 | 0.237996 | 0.505757 |
| sigma | 0.240657 | 0.214807 | 0.271578 |
| theta[1] | 0.712026 | 0.359498 | 0.981214 |
| theta[2] | 0.541167 | 0.382518 | 0.70863 |
| theta[3] | 0.036998 | 0.00097 | 0.139073 |

**Supplementary Table 9: Parameter estimates for the 2 severity group, bivariate antibody kinetics models.**

| **Abbott ARCHITECT & Roche Elecsys (2 groups)** | | | |
| --- | --- | --- | --- |
| **parameter** | **posterior mean** | **2.5%** | **97.5%** |
| lambda_Abbott_ | 0.008933 | 0.007822 | 0.010019 |
| lambda_Roche_ | -0.0021 | -0.00398 | -0.00012 |
| beta_Abbott_[1] | -0.15124 | -0.92086 | 0.526788 |
| beta_Abbott_[2] | 2.197336 | 2.075352 | 2.318977 |
| beta_Roche_[1] | 1.702817 | 0.952109 | 2.39288 |
| beta_Roche_[2] | 4.56304 | 4.361948 | 4.767599 |
| tau_Abbott_[1] | 1.808788 | 1.447634 | 2.238585 |
| tau_Abbott_[2] | 0.32436 | 0.236705 | 0.416222 |
| tau_Roche_[1] | 2.097119 | 1.735337 | 2.513518 |
| tau_Roche_[2] | 0.27835 | 0.096709 | 0.470478 |
| Sigma[1,1] | 0.051496 | 0.040006 | 0.066726 |
| Sigma[1,2] = Sigma[2,1] | 0.036294 | 0.02064 | 0.056062 |
| Sigma[2,2] | 0.143408 | 0.109671 | 0.183508 |
| theta_Abbott_[1] | 0.430924 | 0.295767 | 0.580126 |
| theta_Abbott_[2] | 0.035555 | 0.000877 | 0.129012 |
| theta_Roche_[1] | 0.574107 | 0.434949 | 0.702815 |
| theta_Roche_[2] | 0.132578 | 0.020836 | 0.321786 |

### Supplementary Figures

**Supplementary Figure 1: Sub-national time series of reported SARS-CoV-2 cases in Italy (21 regions).** The serosurvey dates are shown by the light blue bar (range) and dark blue circle (midpoint).

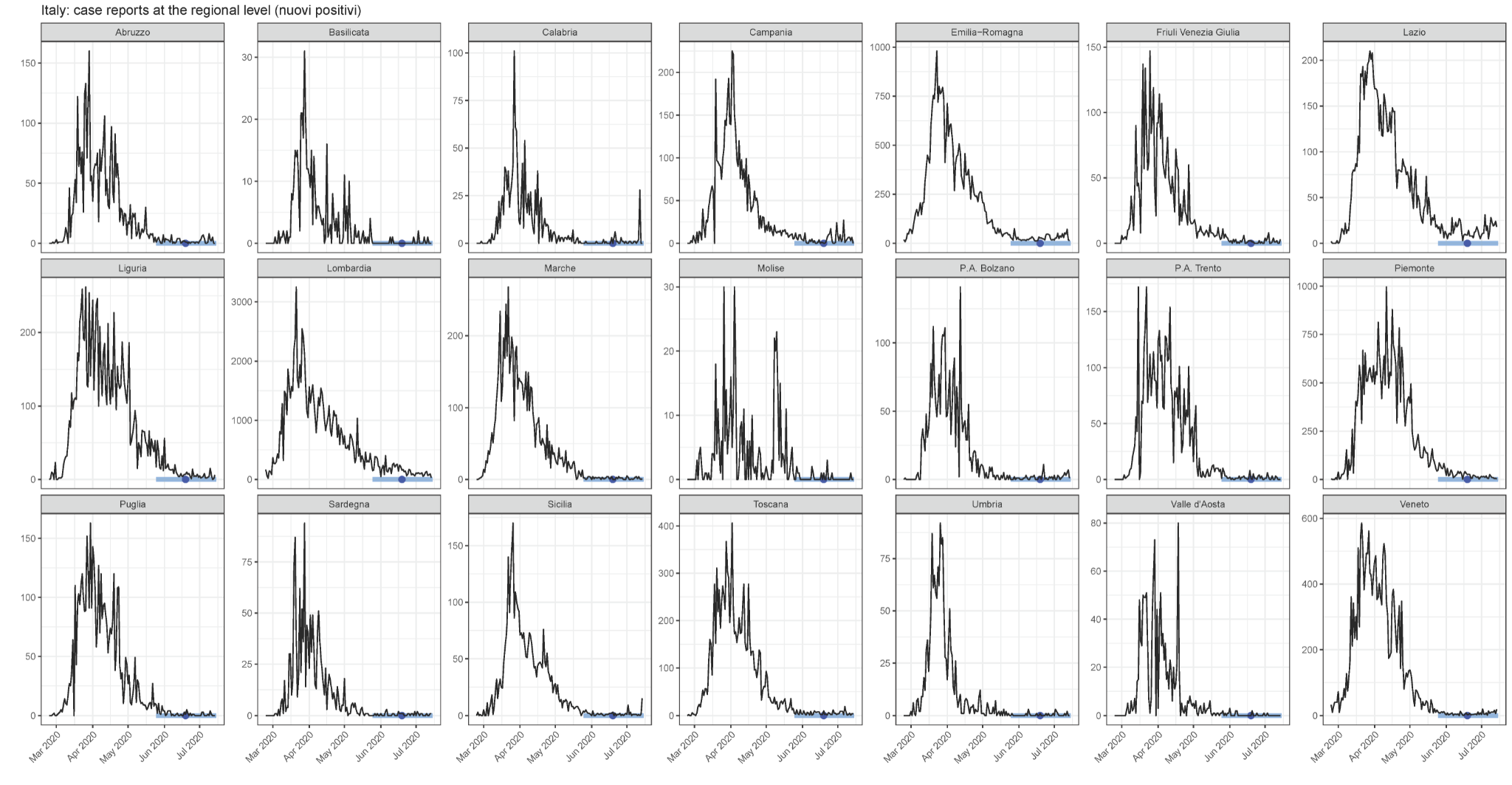

**Supplementary Figure 2: Sub-national time series of reported SARS-CoV-2 cases in Spain (52 provinces).** The serosurvey dates are shown by the light blue bar (range) and dark blue circle (midpoint).

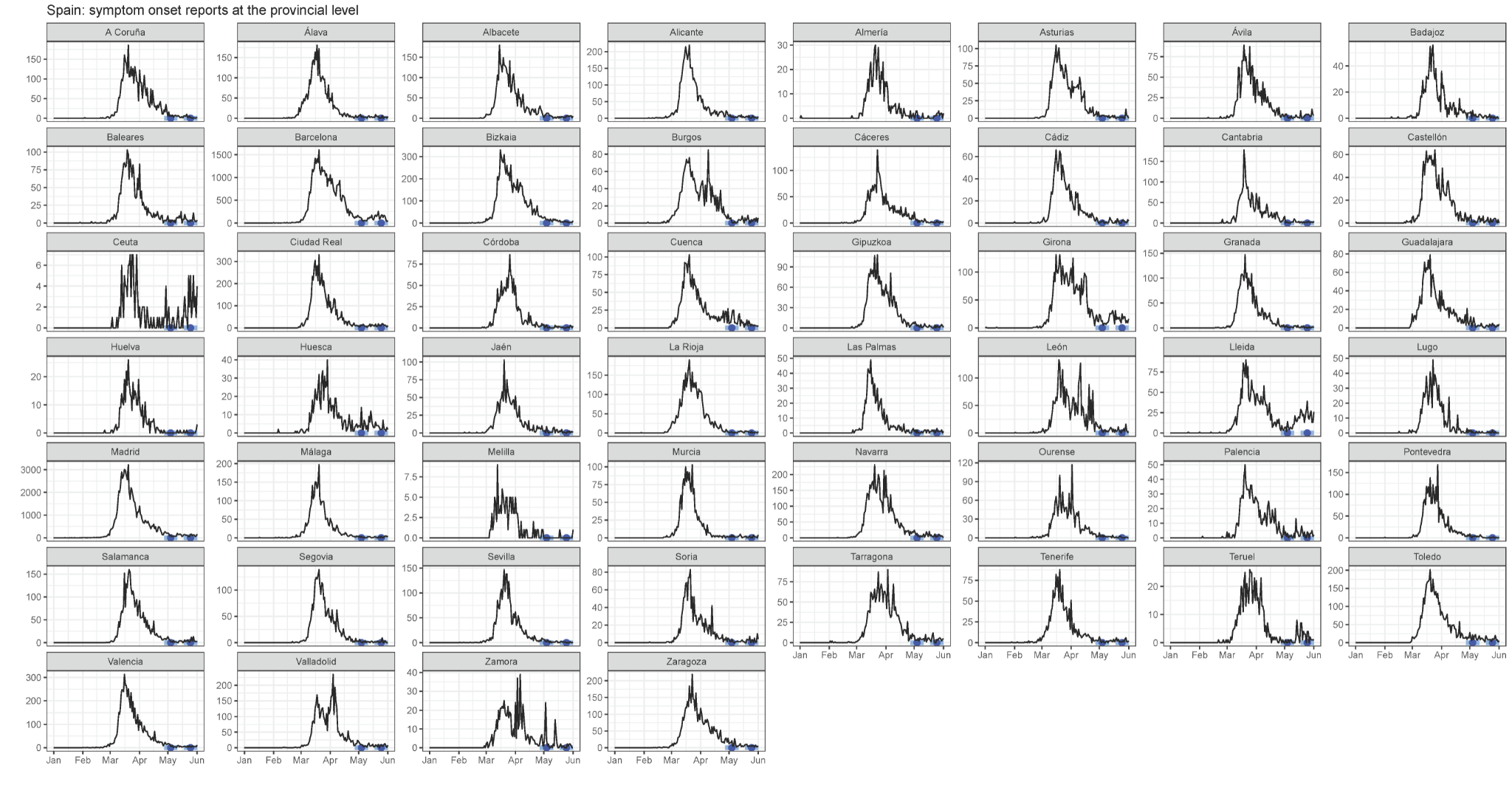

**Supplementary Figure 3: Sub-national time series of reported SARS-CoV-2 cases in the five prefectures of Japan included in this analysis.** The serosurvey dates are shown by the light blue bar (range) and dark blue circle (midpoint).

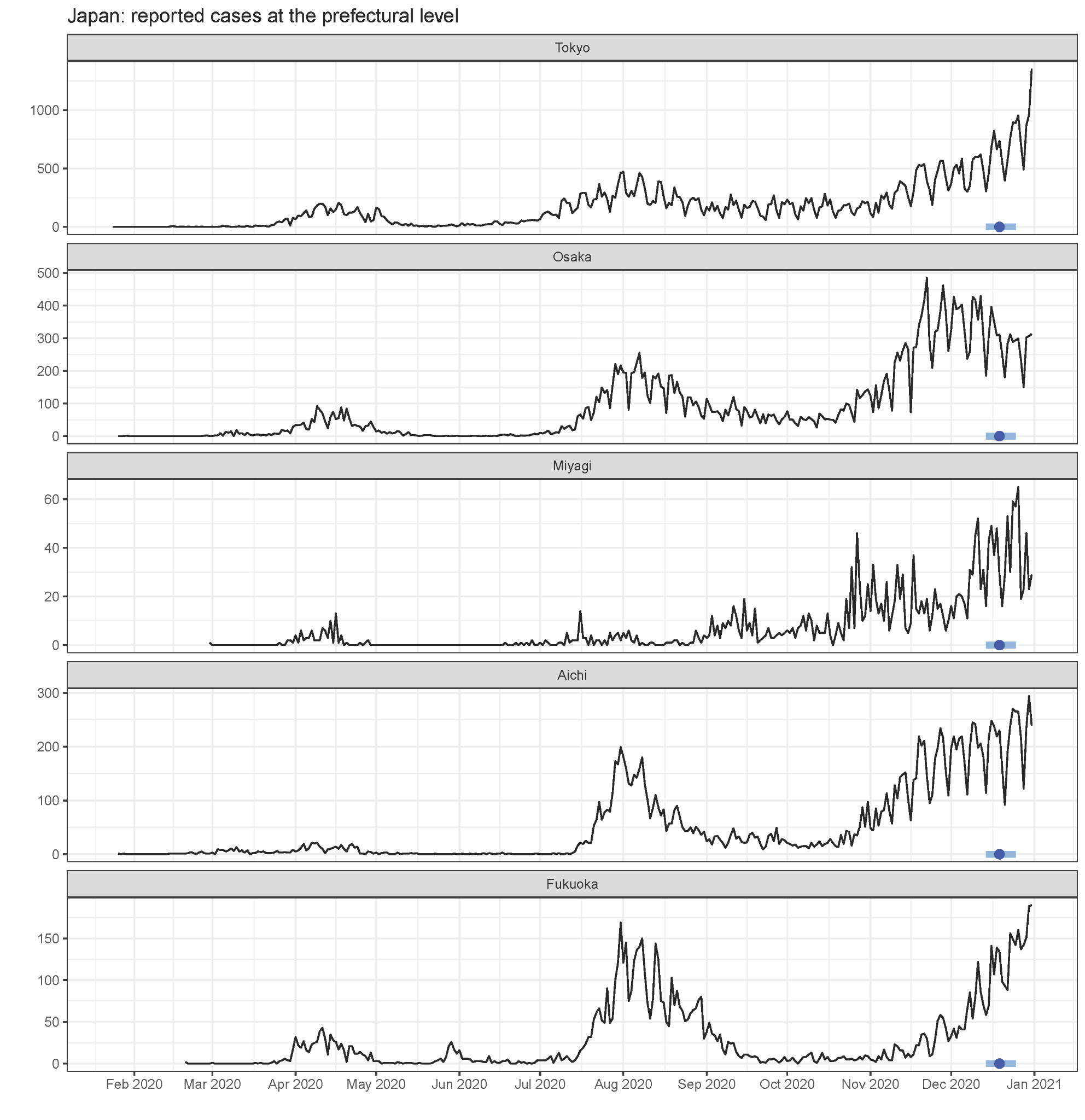

**Supplementary Figure 4: Composition of state-level reconstructed daily time series of symptom onsets in the United States using death reports and the EpiNow2 software, within each census division.**

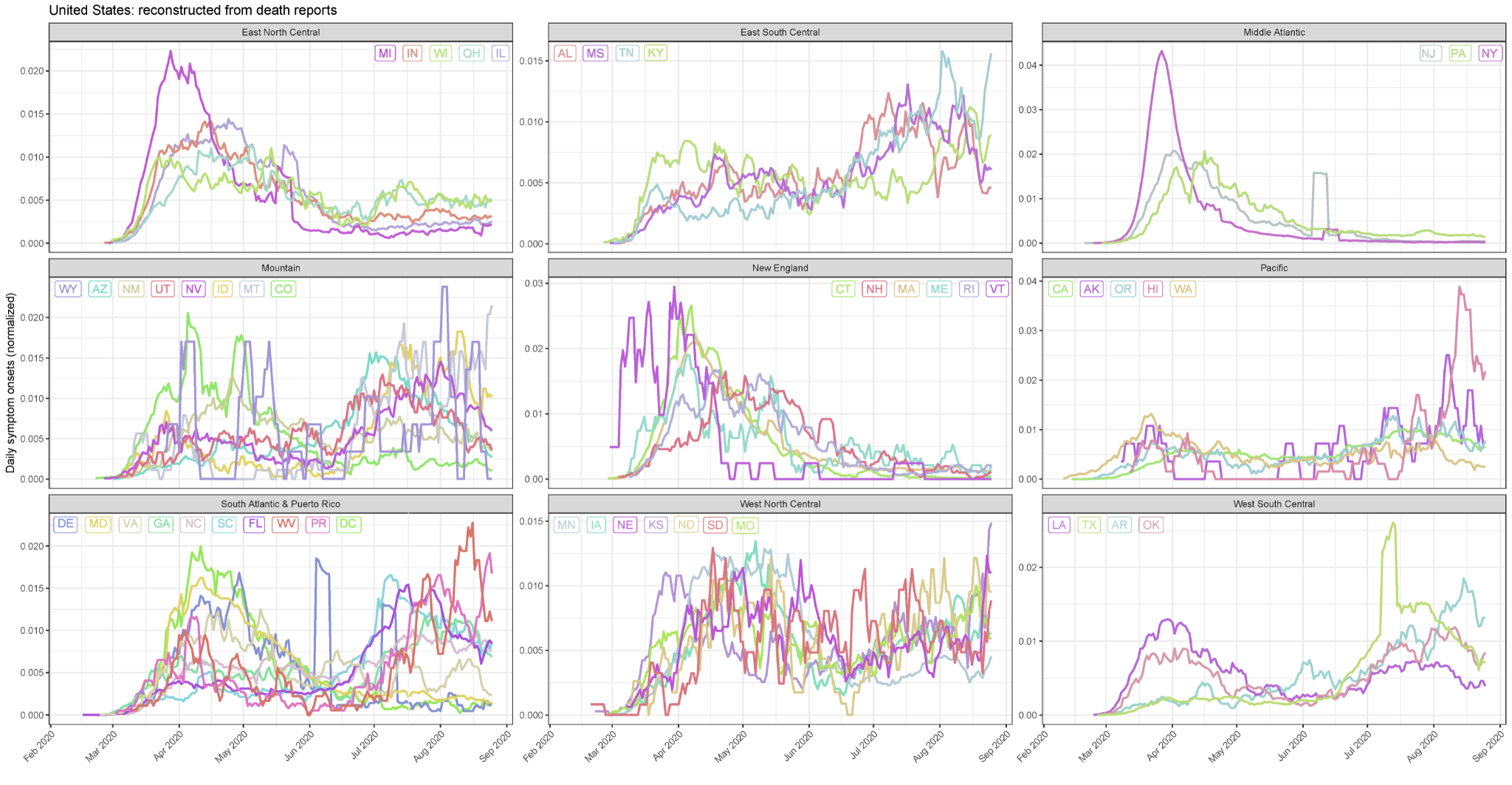

**Supplementary Figure 5: Comparison of reconstructed daily time series of symptom onsets using case reports (red) and death reports (turquoise) and the EpiNow2 software in (A) the United States and in (B) Japan.** Values are normalized to the total case or death reports in the locale for comparability. Posterior median estimates are shown as the solid line and the 95% credible intervals in the shaded area. The serosurvey dates are shown by the light blue bar (range) and dark blue circle (midpoint). These reconstructed symptom onsets, while different in absolute magnitude, were relatively consistent within a population.

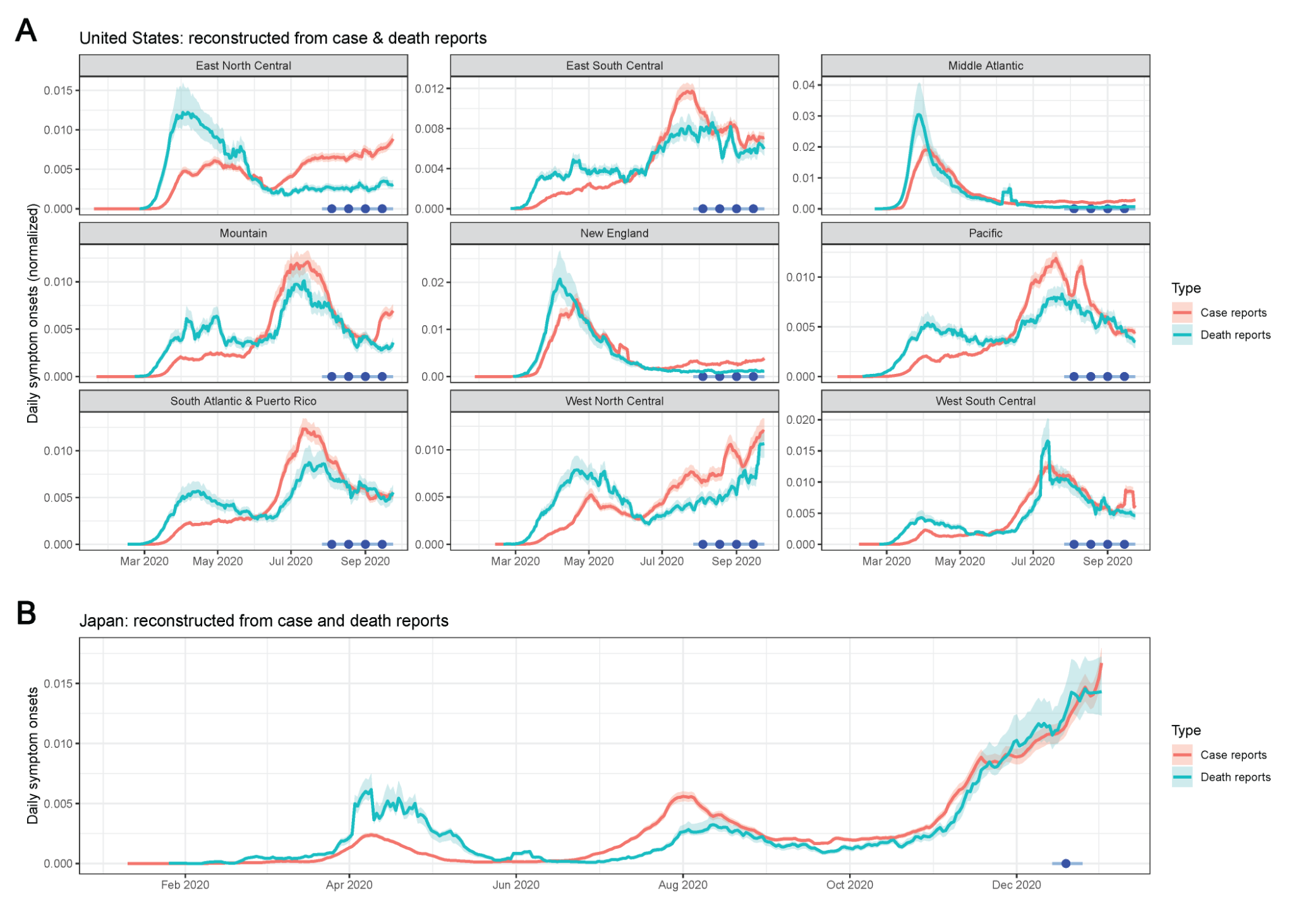

**Supplementary Figure 6: Comparison of reconstructed daily time series of symptom onsets in Manaus, Brazil using hospitalization reports.** (*Upper*) Using time delay distribution from Bi *et al* [[11]](https://paperpile.com/c/CEaFMo/whVd). (*Lower*) Using time delay distribution from Linton *et al* [[12]](https://paperpile.com/c/CEaFMo/nFtz) (used in main text). See Supplementary Table 3 for values.

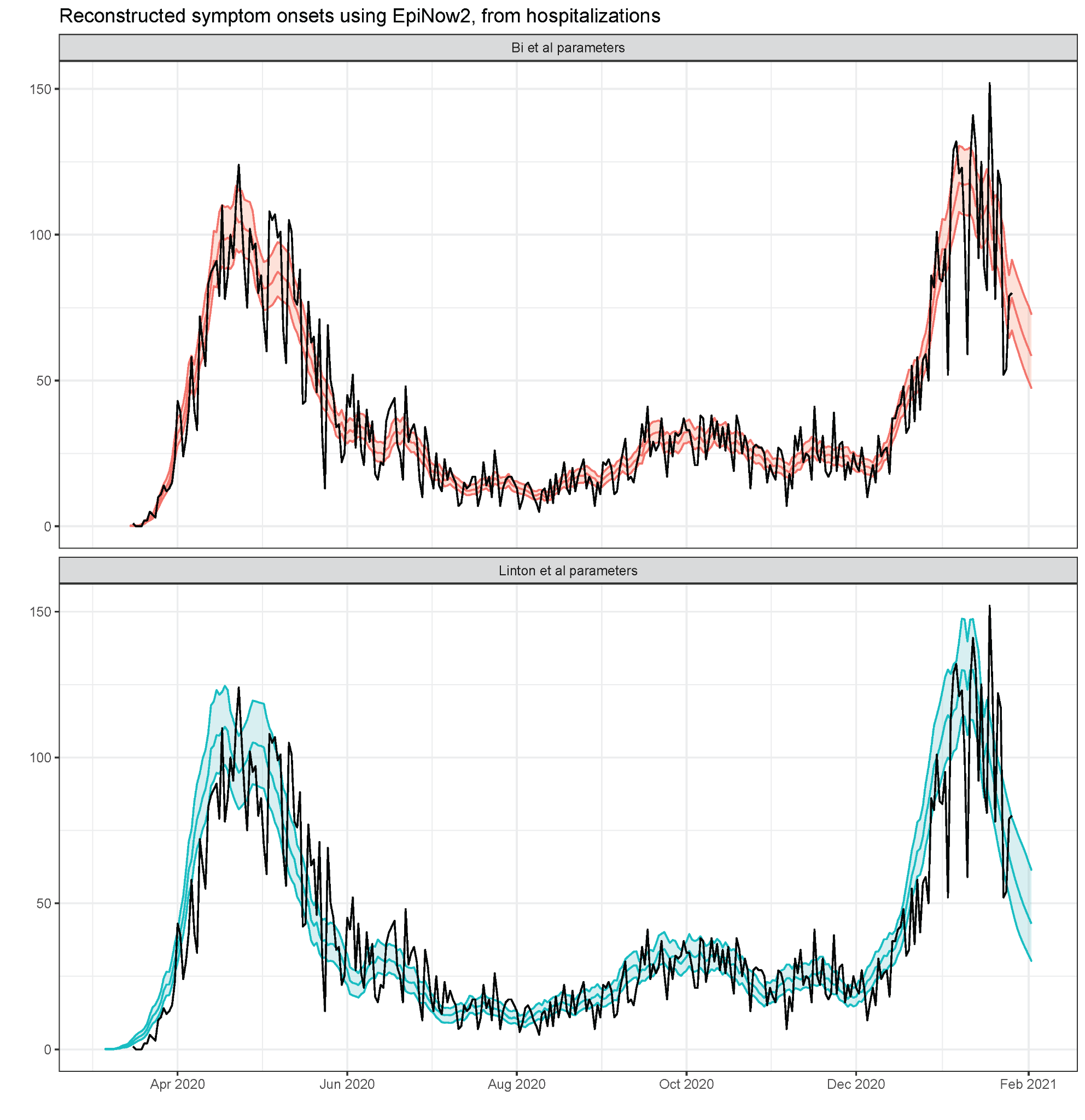

**Supplementary Figure 7: Interpolated age-specific disease severities in Manaus, Brazil. (A)** Age-specific probabilities of hospitalization and **(B)** age-specific probabilities of experiencing symptoms, conditional on SARS-CoV-2 infection, for the age bins in Supplementary Table 5 (*upper row*) and interpolated for the age bins reported in the serosurvey data from Manaus (*lower row*). Age-specific hospitalization probabilities are based on values published in [[21]](https://paperpile.com/c/CEaFMo/YFAr), and age-specific symptom probabilities are based on values published in [[22]](https://paperpile.com/c/CEaFMo/VczA).

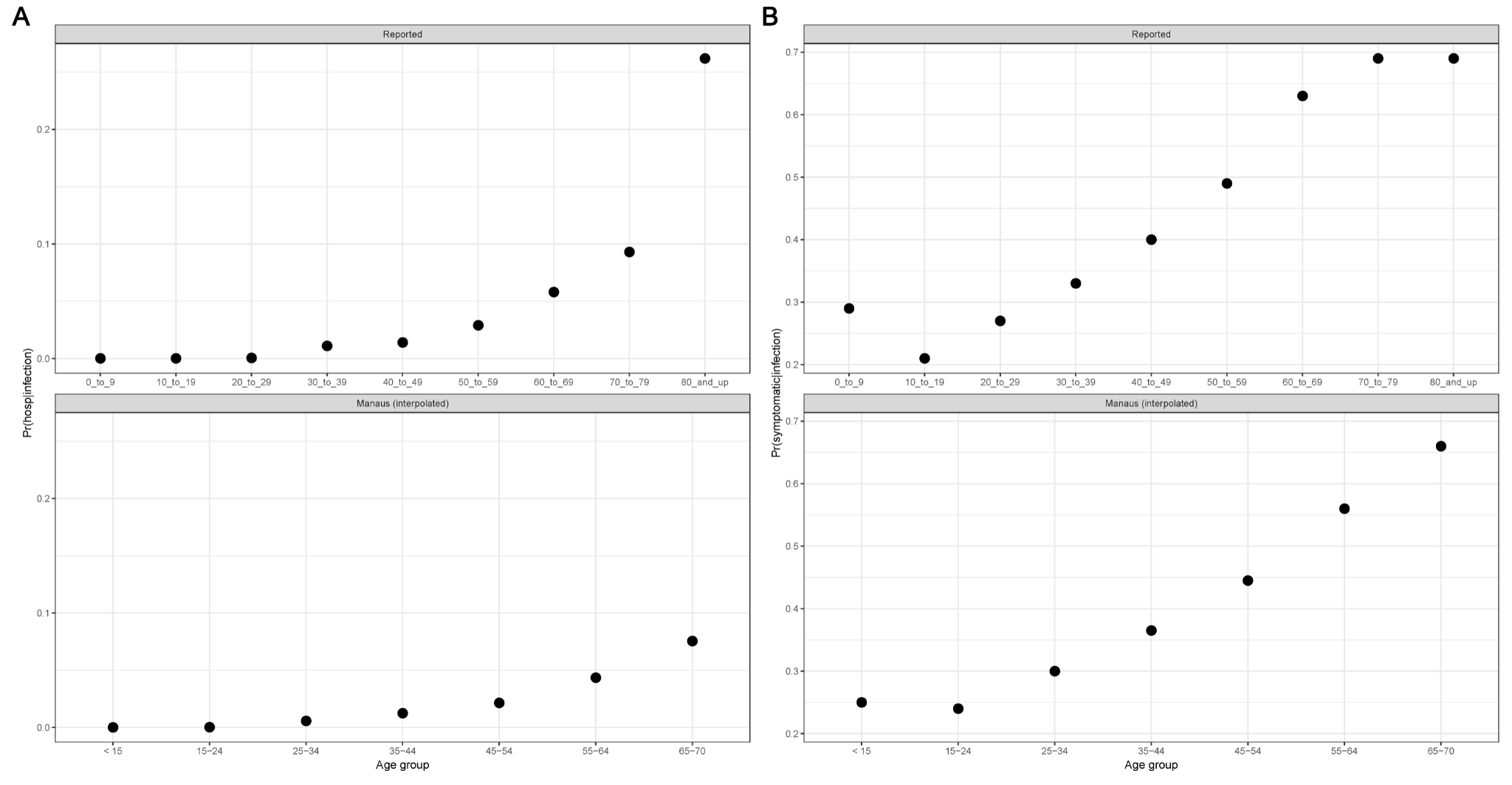

**Supplementary Figure 8: Additional data on long-term longitudinal kinetics by assay.** Additional longitudinal samples from 7 asymptomatic individuals were tested on each assay. Data in orange were included for fitting the antibody kinetics models, and the fitted values for each individual over time are shown. Black stars represent the new data. Red dashed line indicates the cutoff value for positivity.

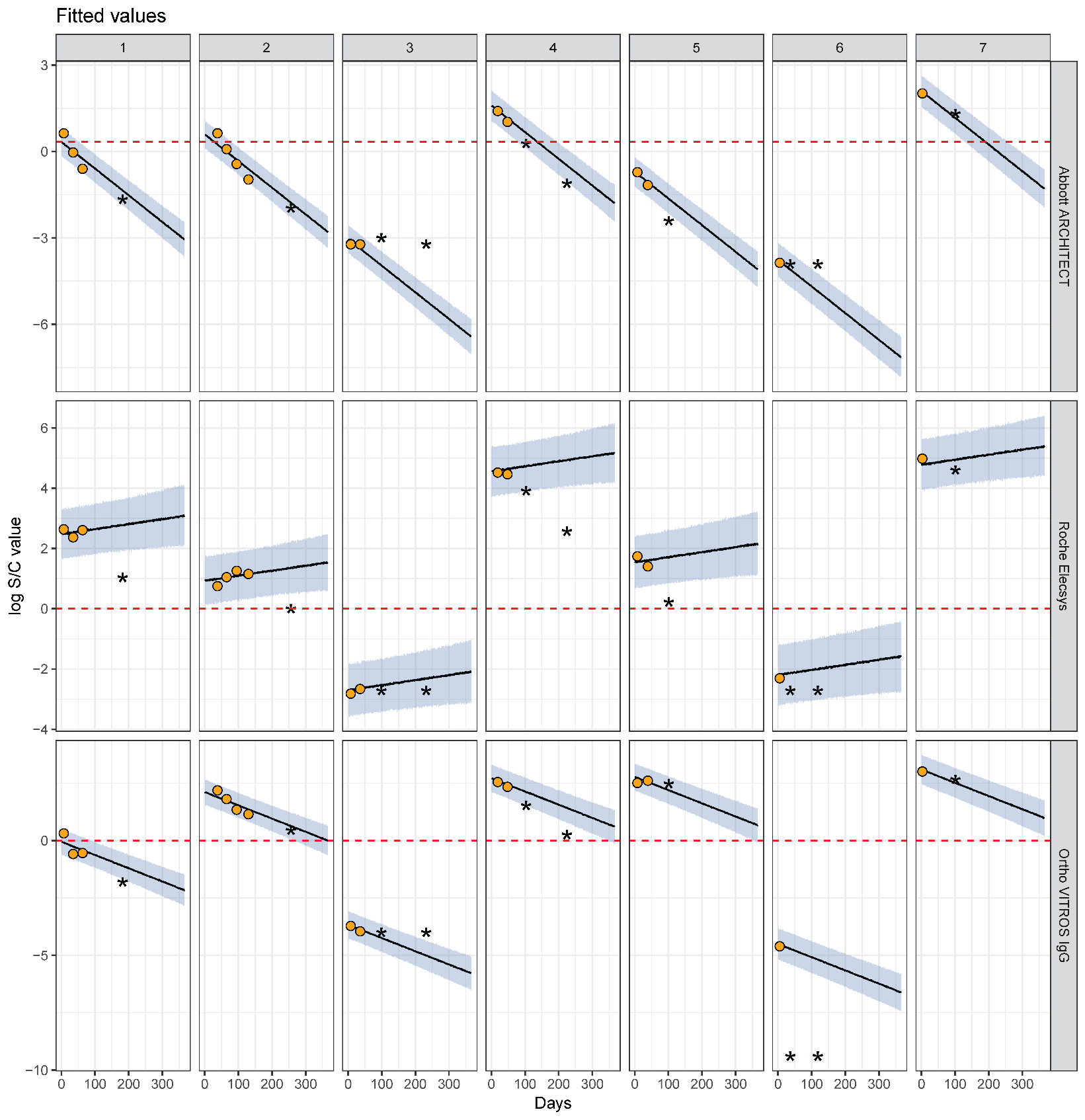

**Supplementary Figure 9: Longitudinal SARS-CoV-2 antibody kinetics and estimated assay sensitivities by time and disease severity (3 severity groups).** This figure is analogous to Figure 3 in the main text, except now further partitioning the non-hospitalized group into asymptomatic and symptomatic, non-hospitalized individuals. (*Upper row*) Time since symptom onset (offset by 3 weeks) is shown on the x-axis versus the log-transformed antibody response for each of the Abbott ARCHITECT, Roche Elecsys, and Ortho VITROS IgG assays, stratified by disease severity. For asymptomatic individuals, the time since the first positive PCR test (offset by 3 weeks) was used. This time-metric is referred to as ‘time since seroconversion’ hereafter. Longitudinal samples are connected by black lines. Black dotted lines indicate cutoff values for positivity on that assay. (*Lower row*) Estimated sensitivity of each assay (showing posterior median estimates as the solid line and 95% credible intervals), stratified by disease severity, from 0 to 365 days after seroconversion. The dashed vertical line in purple indicates the maximum observed time on the corresponding panel above (i.e., x=130 for asymptomatic, x=136 for symptomatic and non-hospitalized, and x=118 for hospitalized).

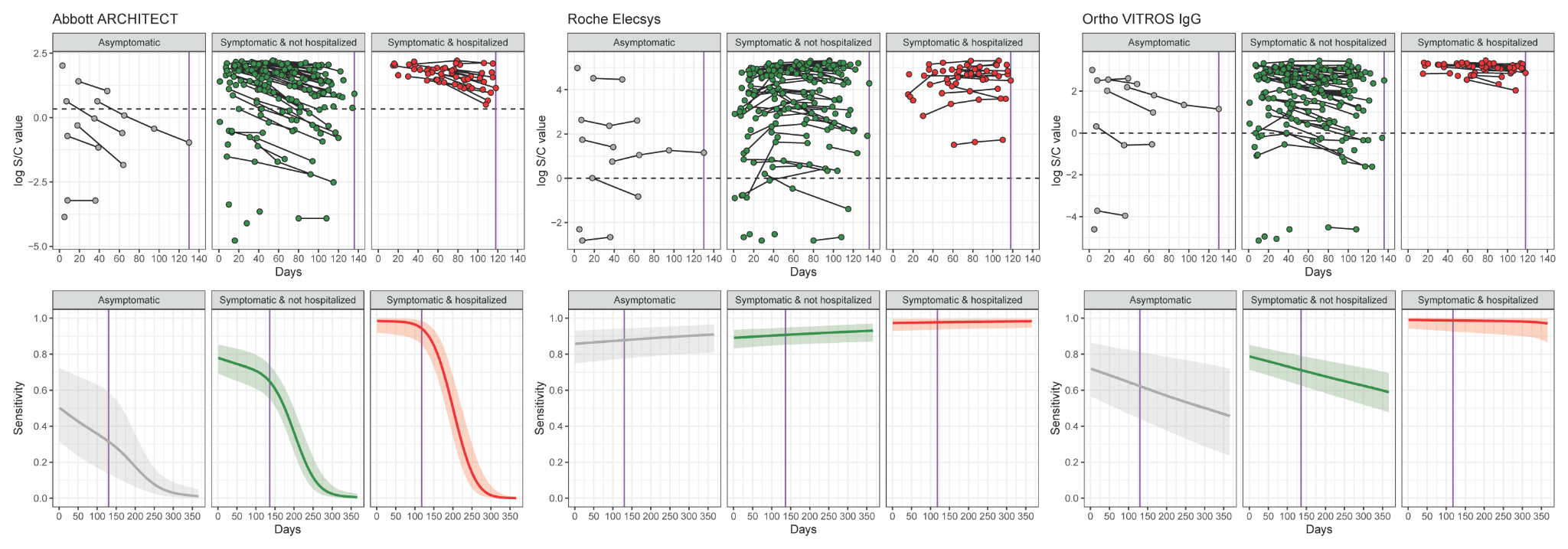

**Supplementary Figure 10: Comparison of kinetics data from Manaus, Brazil to kinetics data from the LIINC cohort. (A)** Longitudinal antibody kinetics in donors from Manaus with a positive antibody response on the Abbott ARCHITECT assay (cutoff: S/C value = 1.4), stratified by the month of first positive donation between March and August 2020. **(B)** Estimated slope ($\lambda$) from a random effects regression for each month in Manaus, compared to the estimated slope for this assay from antibody kinetics in the LIINC cohort with 2 severity groups (purple rectangle).

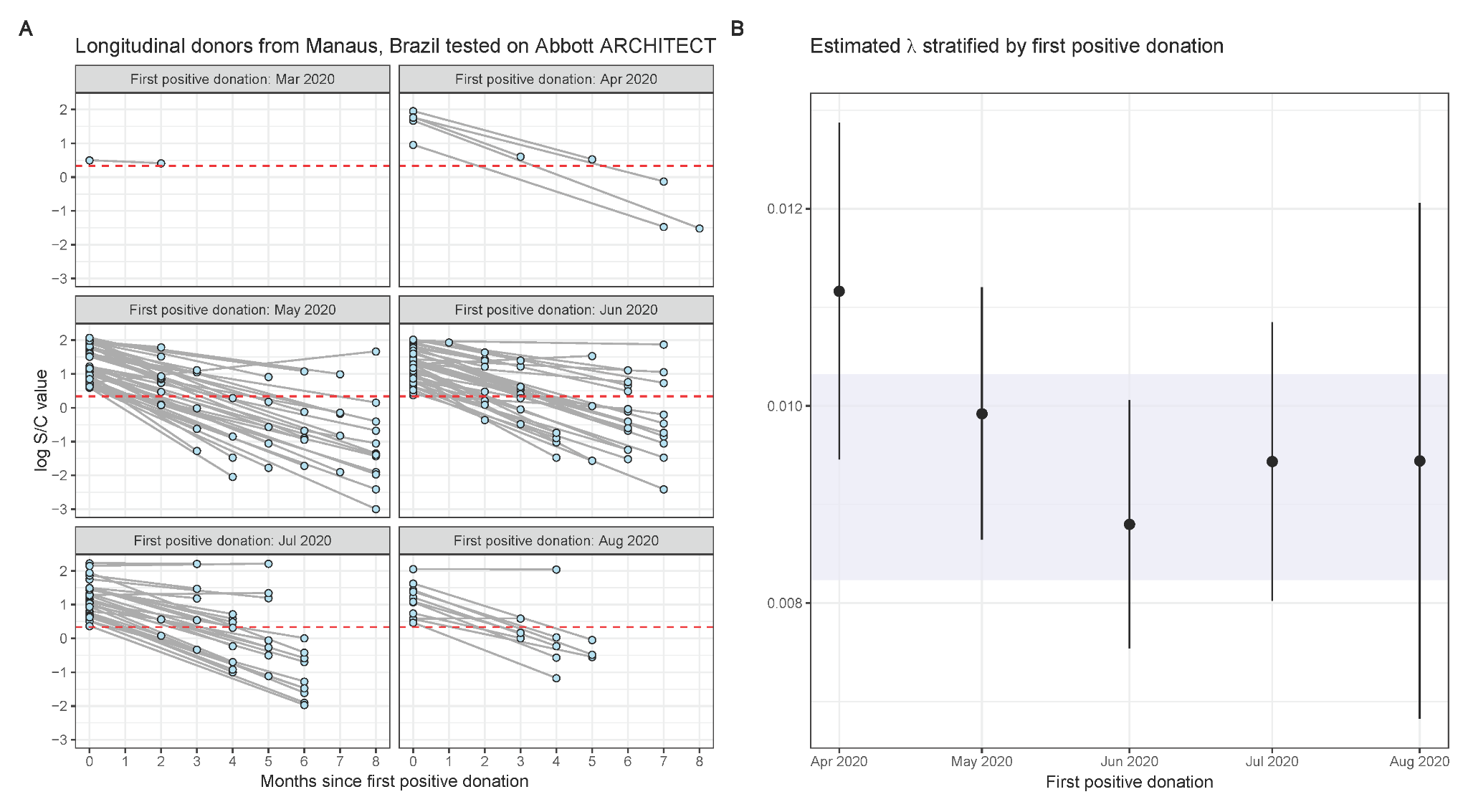

**Supplementary Figure 11: Estimated bivariate sensitivity for the Abbott ARCHITECT and Roche Elecsys assays (showing posterior median estimates as the solid line and 95% credible intervals), stratified by hospitalization status, from 0 to 365 days after seroconversion.** Estimates under the 2 severity group scenario.

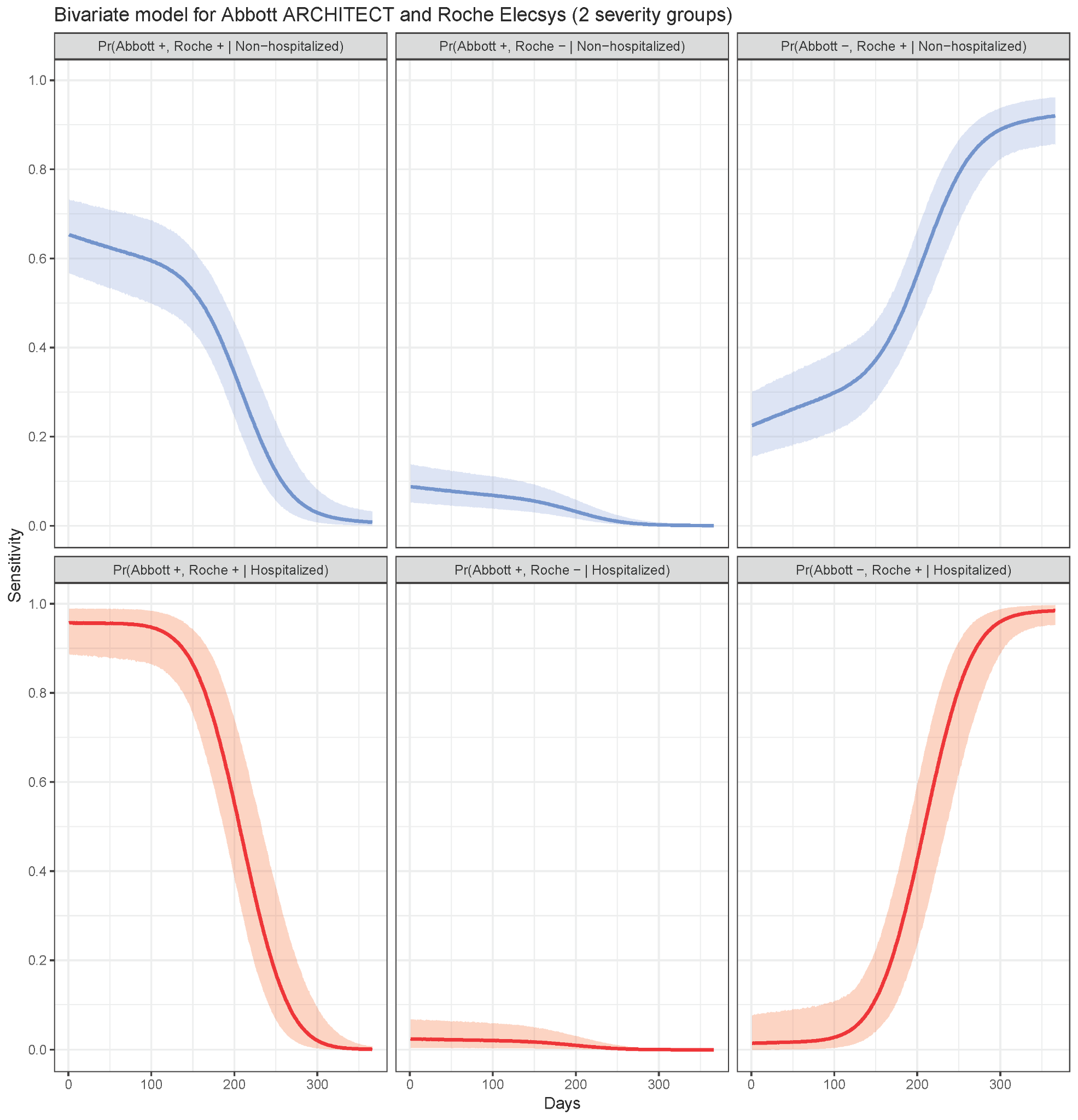

**Supplementary Figure 12: Relative bias in seroprevalence estimation (3 severity groups).** This figure is analogous to Figure 5 in the main text, except now further partitioning the non-hospitalized group into asymptomatic and symptomatic, non-hospitalized individuals. For each panel, the raw seroprevalence result is shown on the x-axis and the ratio of the adjusted to raw seroprevalence is shown on the y-axis (median and 95% credible interval). The ratio equaling 1 (i.e., no bias) is shown in the dashed line. **(A)** Italy, where each point represents a region. **(B)** Spain, for (*upper*) Round 1 and (*lower*) Round 2, where each point represents a province. **(C)** The 9 census divisions of the United States, where the color of the point represents the survey round. **(D)** Manaus, Brazil, where each point represents a month. As in Figure 4 in the main text, for panels C and D, the adjusted seroprevalence estimates are weighted by population demography and age-specific disease severity. **(E)** Japan, where each point represents a prefecture. The scenario considered here is the case of using the results of the two assays.

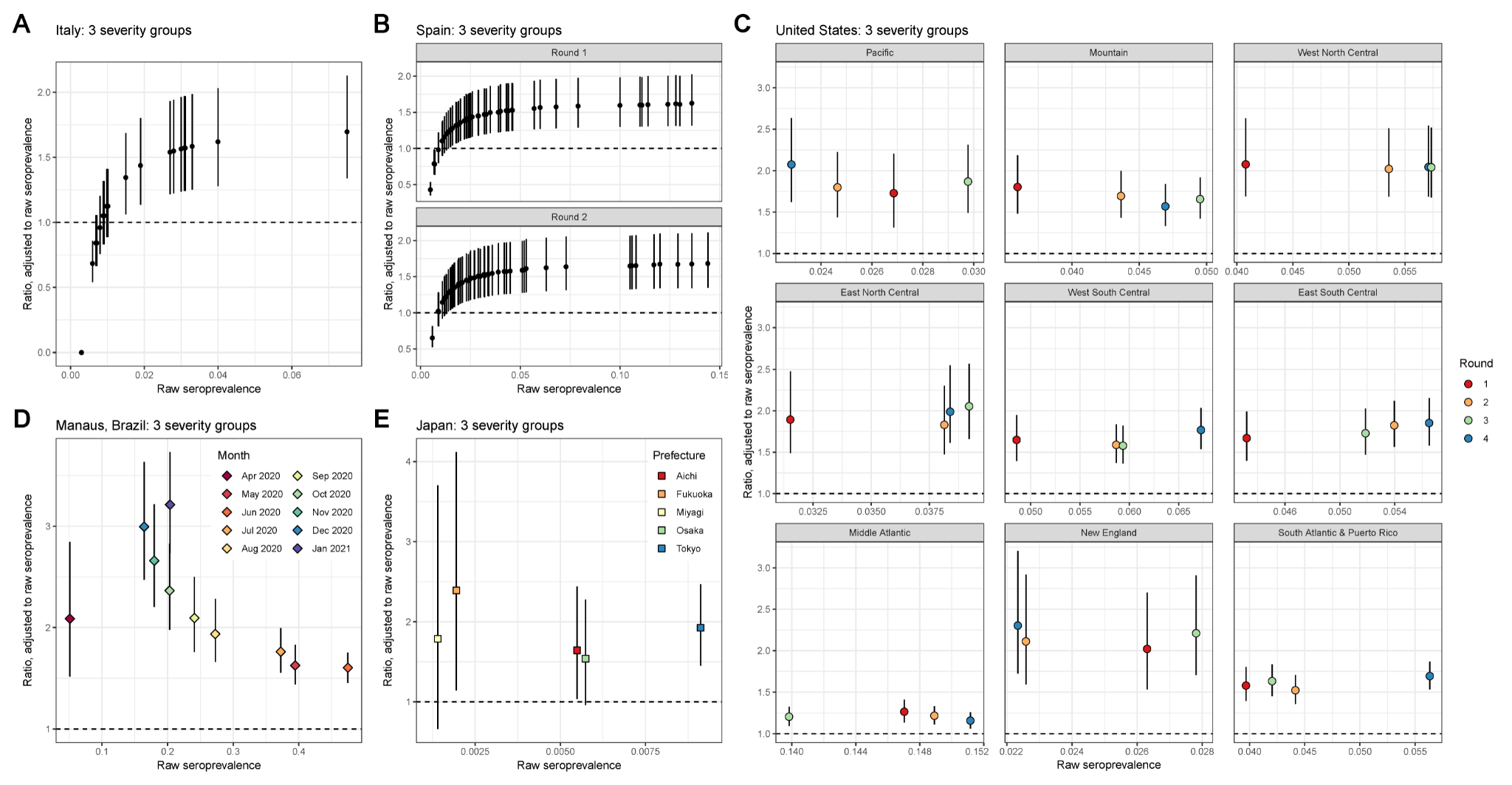

**Supplementary Figure 13: Posterior median seroprevalence and 95% credible intervals (CrI) for Italy by region, under the 2 and 3 severity group scenarios.**

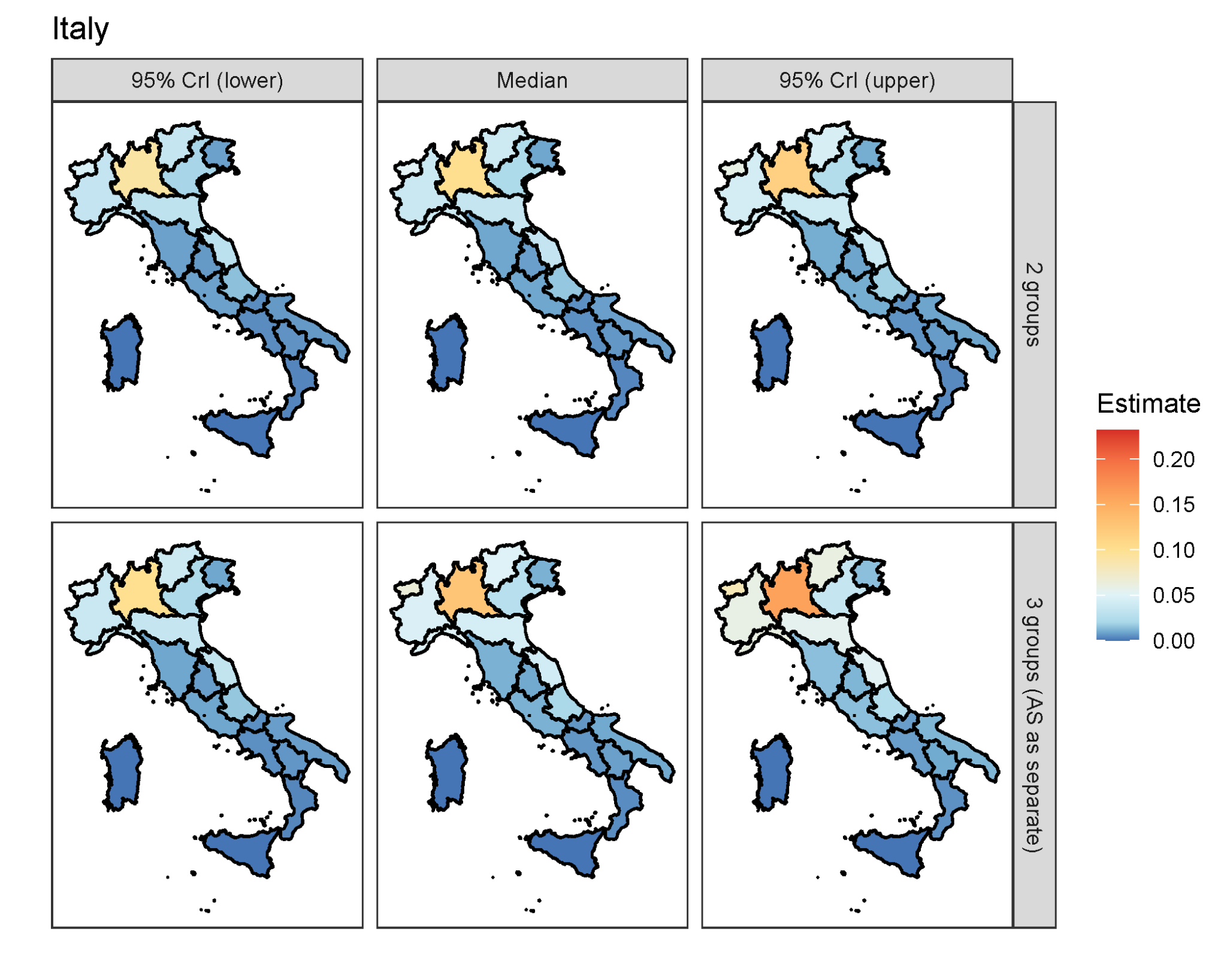

**Supplementary Figure 14: Posterior median seroprevalence and 95% credible intervals (CrI) for the two rounds in Spain by province, under the 2 and 3 severity group scenarios.**

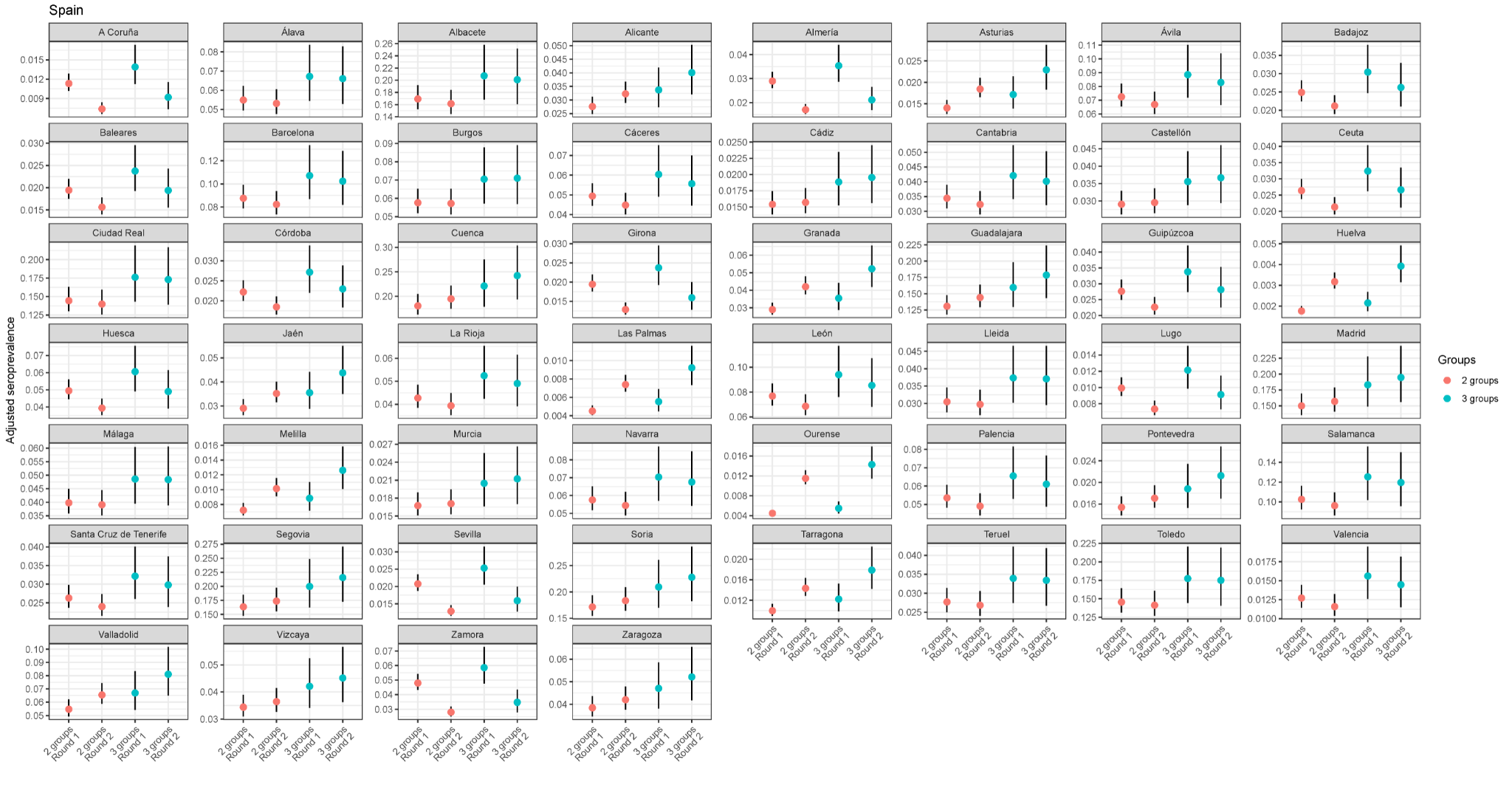

**Supplementary Figure 15: Distribution of the numbers of samples tested in US CDC serosurvey by census division, state, and round.** These data are available at [[24]](https://paperpile.com/c/CEaFMo/7G0A).

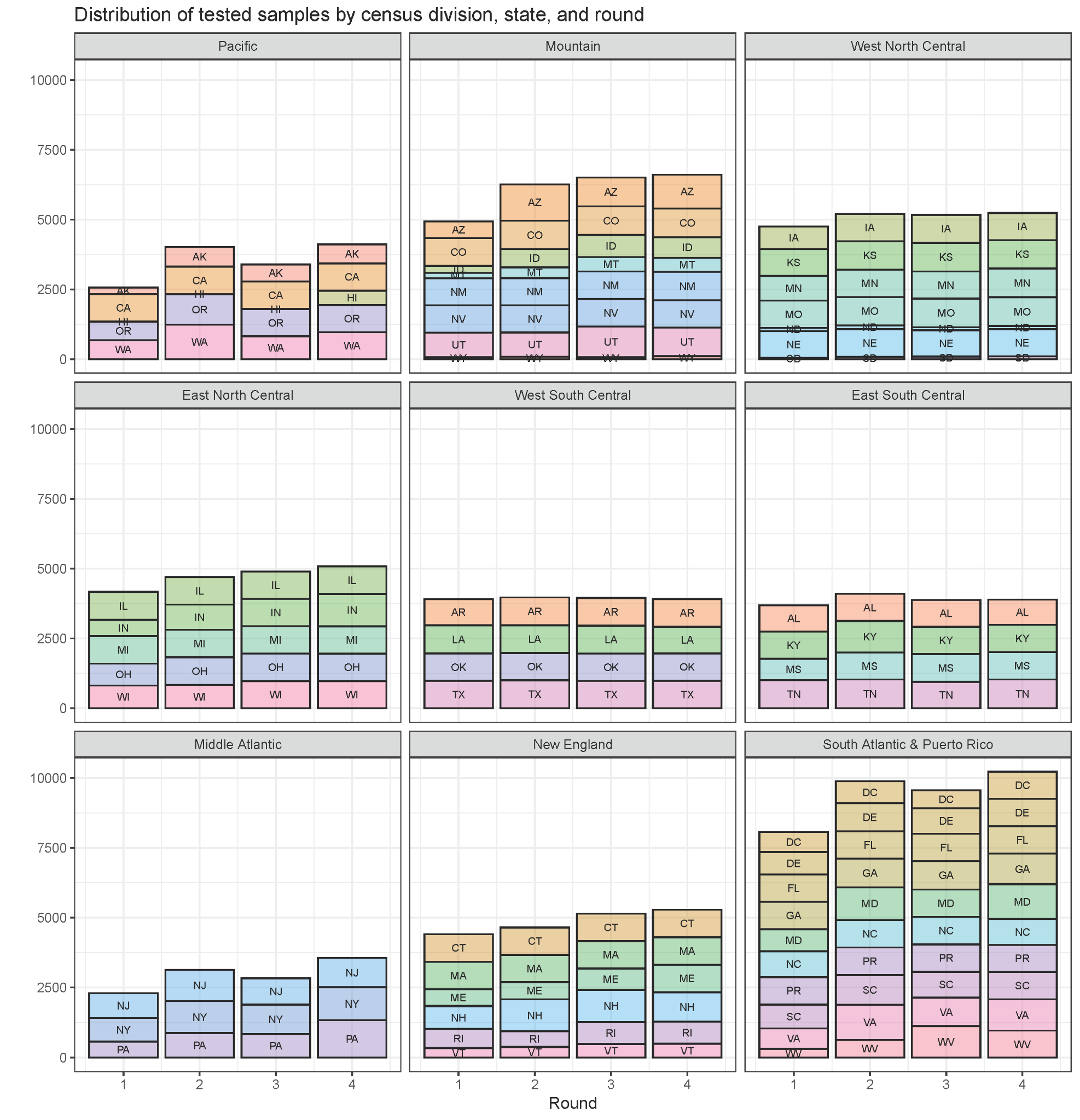

**Supplementary Figure 16: Distribution of the numbers of samples tested in the US CDC serosurvey by state, age group, and round.** These data are available at [[24]](https://paperpile.com/c/CEaFMo/7G0A).

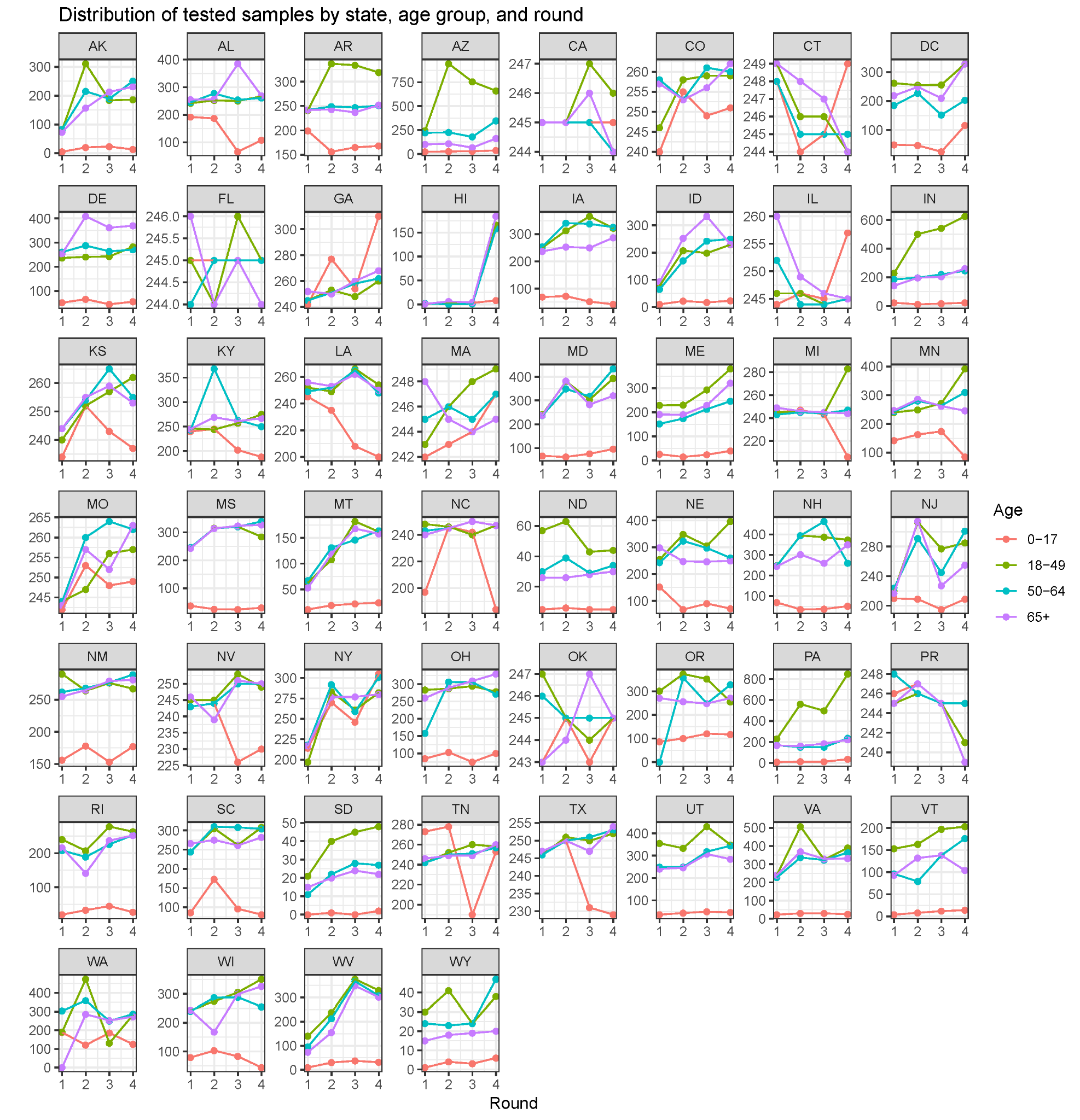

**Supplementary Figure 17: Distribution of the numbers of samples tested in the US CDC serosurvey by census division, age, sex, assay, and round.**

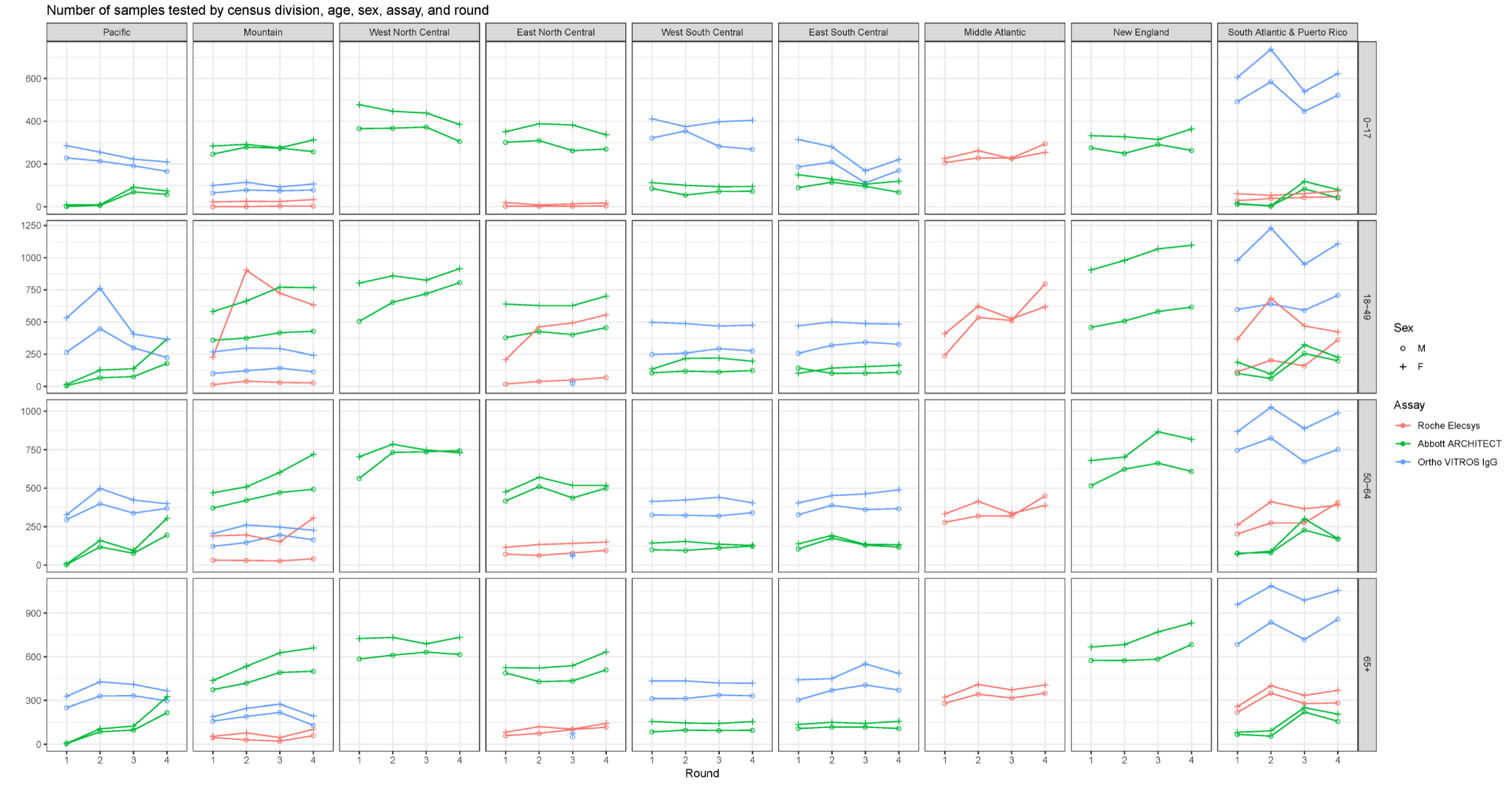

**Supplementary Figure 18: Distribution of the proportions of samples testing positive in the US CDC serosurvey by census division, age, sex, assay, and round.**

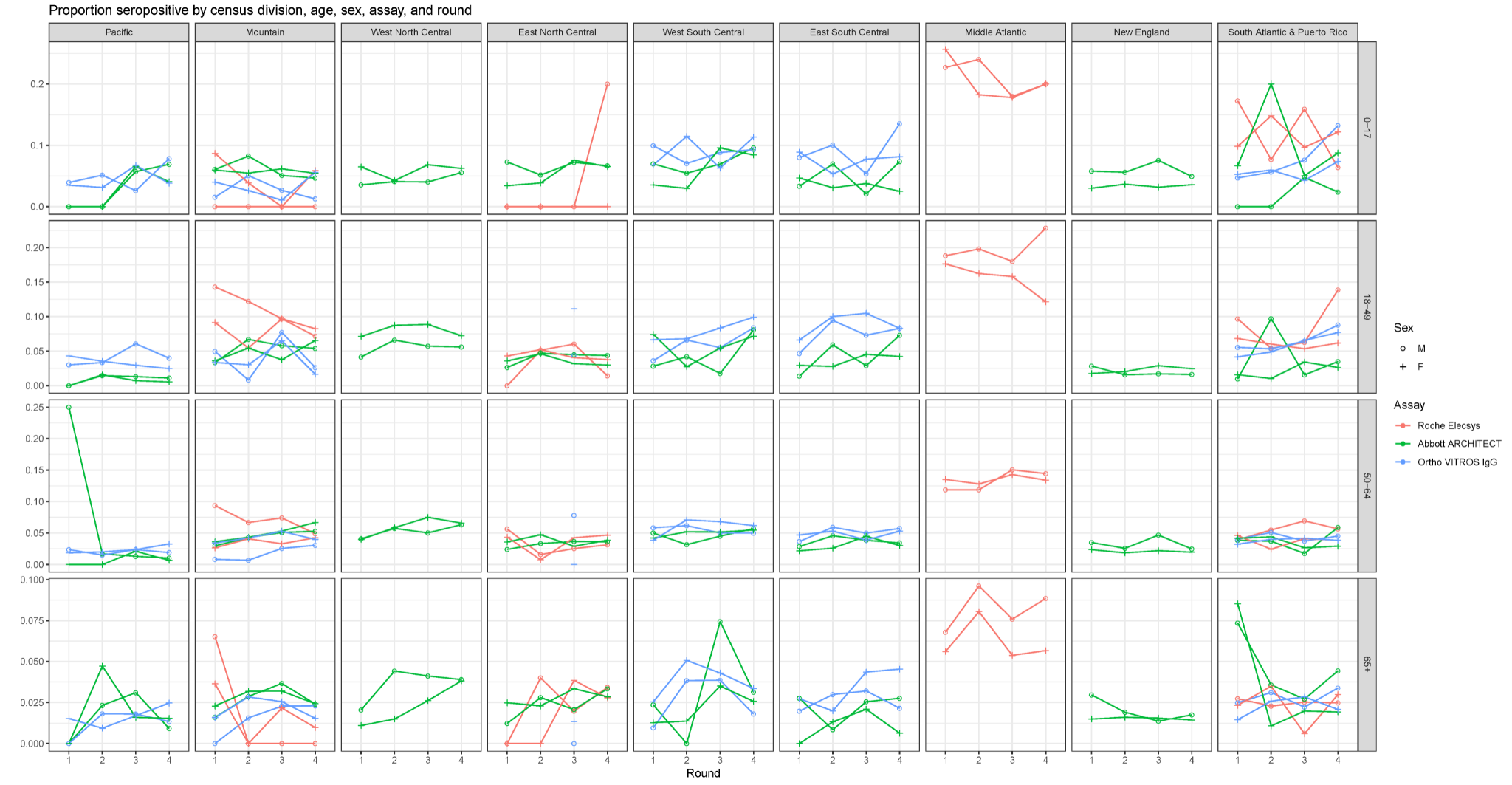

**Supplementary Figure 19: Posterior median seroprevalence by census division in the United States, under the 2 severity group scenario. (A)** Seroprevalence for each of the 4 serosurvey rounds, using symptom onsets reconstructed from death reports (primary scenario). **(B)** Difference in seroprevalence when using symptom onsets reconstructed from case reports, in additive units of seroprevalence.

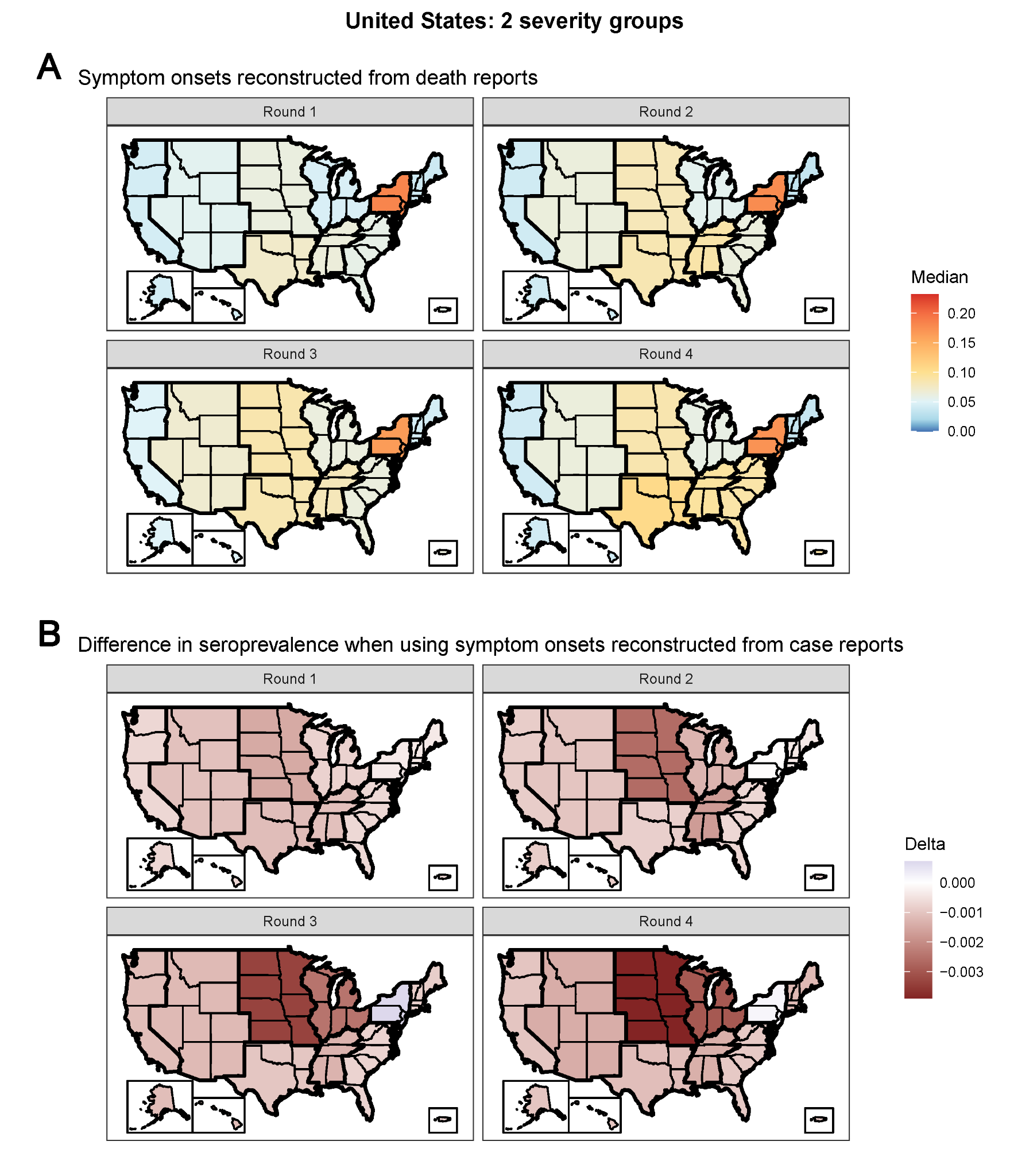

**Supplementary Figure 20: Seroprevalence estimates in the United States under the 2 severity group scenario, stratified by census division, age group, sex, and survey round.** Raw seroprevalence on the x-axis, and posterior median seroprevalence and 95% credible intervals (CrI) on the y-axis. Guides corresponding to the ratio of adjusted to raw seroprevalence equaling 1 (i.e., no bias), 1.5, 2, and 3 depicted by the line types.

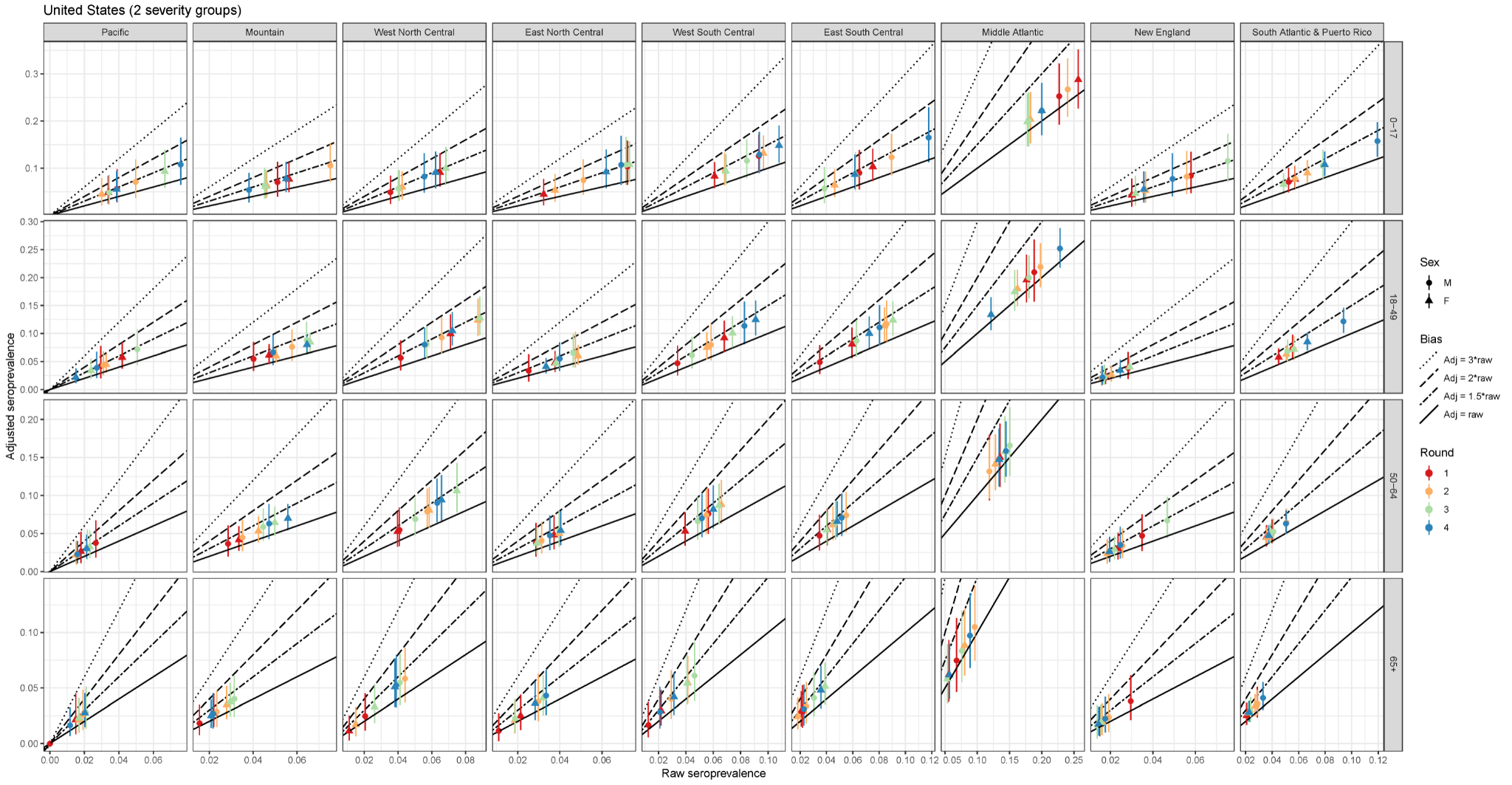

**Supplementary Figure 21: Comparison of seroprevalence estimates in the United States under the 2 severity group scenario, stratified by census division and survey round, to re-calculated estimates from Bajema *et al*.** Seroprevalence from [[13]](https://paperpile.com/c/CEaFMo/XoZk) aggregated to the census division level by state population sizes for each round on the x-axis, and posterior median seroprevalence and 95% credible intervals (CrI) on the y-axis. Guides corresponding to the ratio of adjusted to raw seroprevalence equaling 1 (i.e., no bias), 1.5, and 2 depicted by the line types.

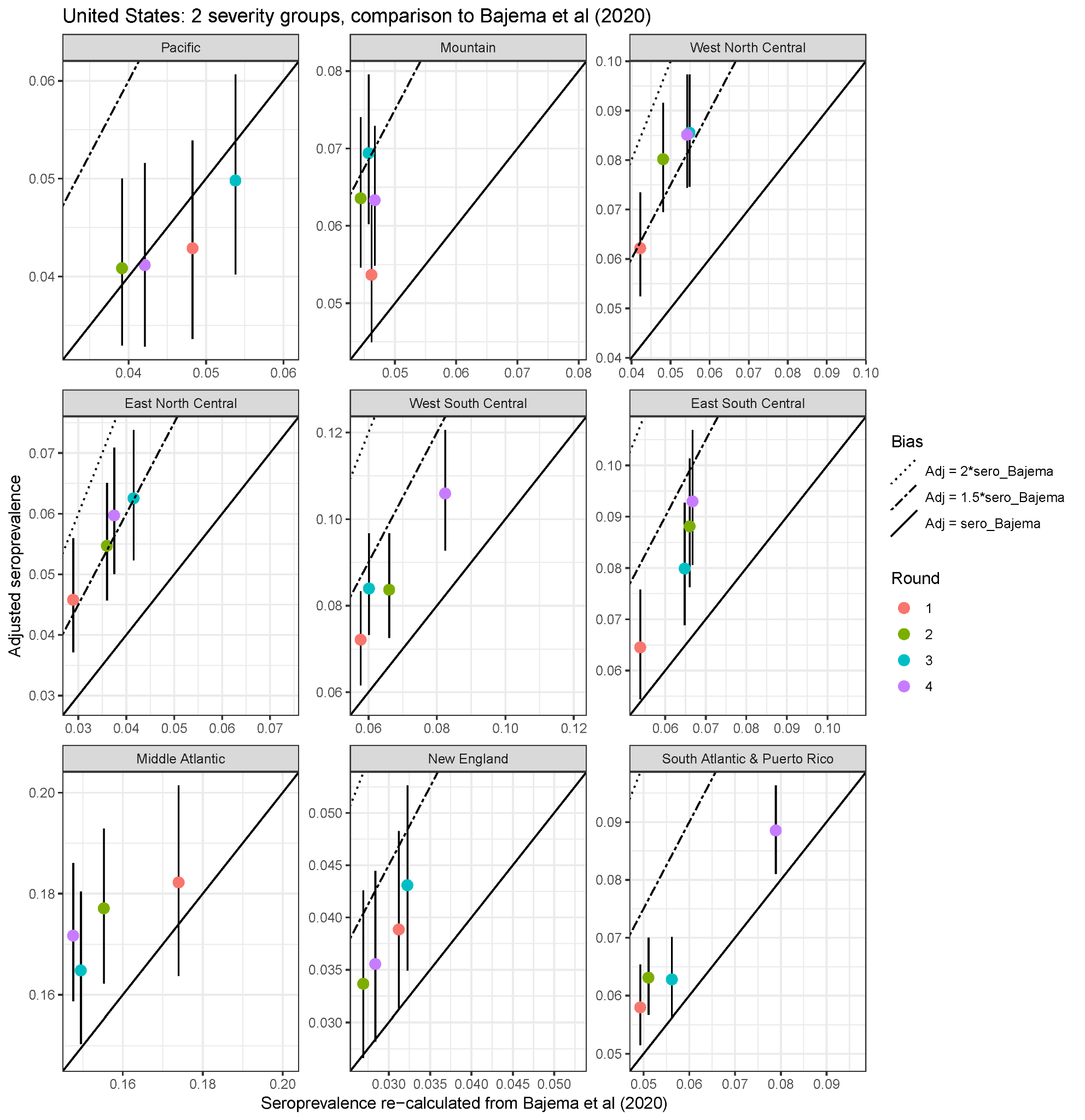

**Supplementary Figure 22: Seroprevalence estimates in Manaus, Brazil under the 2 severity group scenario, stratified by age group, sex, and month.** Raw seroprevalence on the x-axis, and posterior median seroprevalence and 95% credible intervals (CrI) on the y-axis. Guides corresponding to the ratio of adjusted to raw seroprevalence equaling 1 (i.e., no bias), 1.5, 2, and 3 depicted by the line types.

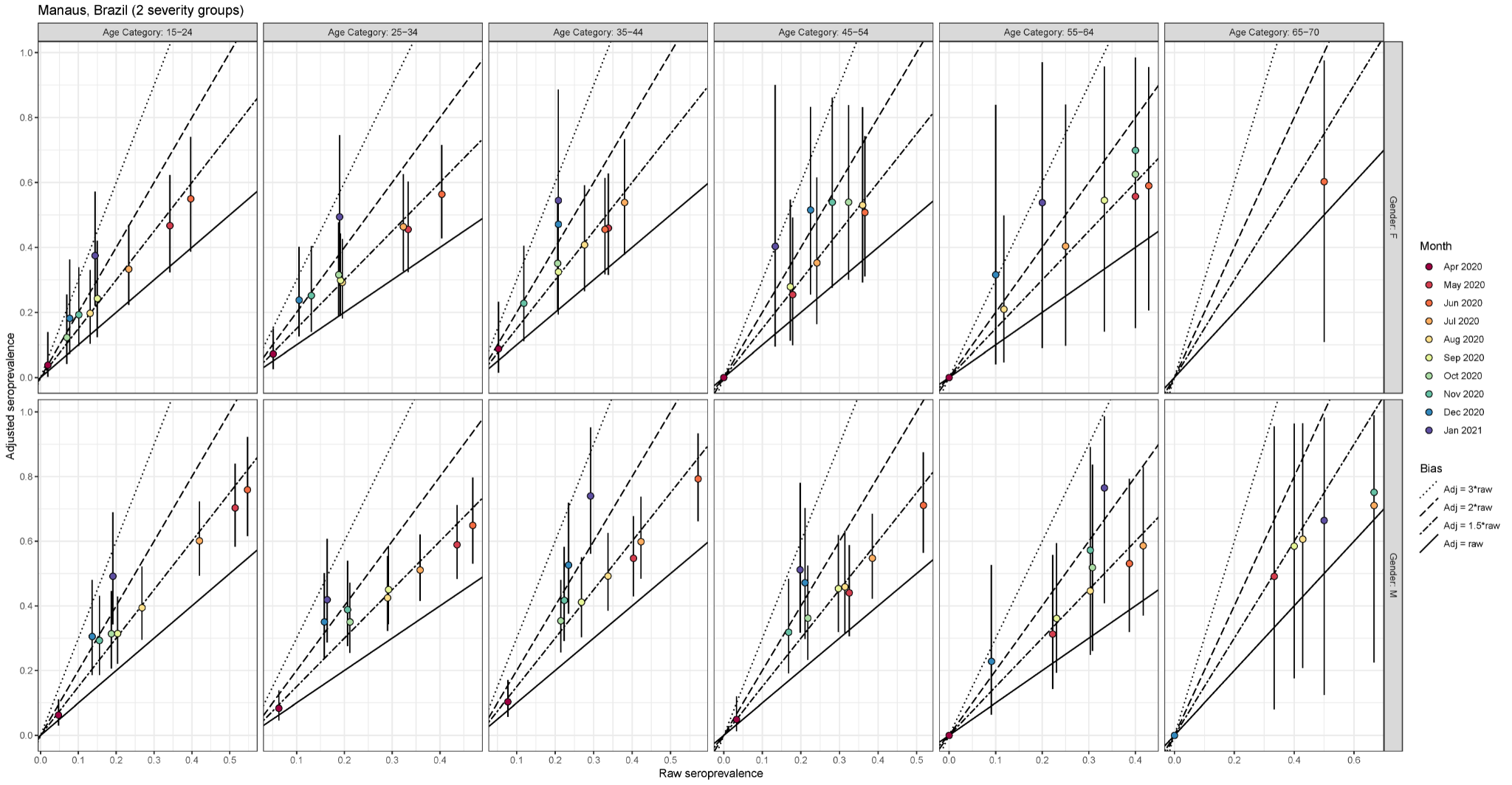

**Supplementary Figure 23: Estimated seroprevalence for the 5 prefectures in Japan, considering the raw results from the (*left*) Abbott ARCHITECT assay only, (*center*) Roche Elecsys assay only, and (*right*) both assays. (A)** Estimates under the 3 severity group scenario, using symptom onsets reconstructed from death reports. **(B)** Estimates under the 2 severity group scenario, using symptom onsets reconstructed from case reports. The raw seropositive proportion is shown in black and the estimated seroprevalence is shown in red. When considering the results from both assays, the raw seropositive proportion is the proportion of samples that tested positive on both.

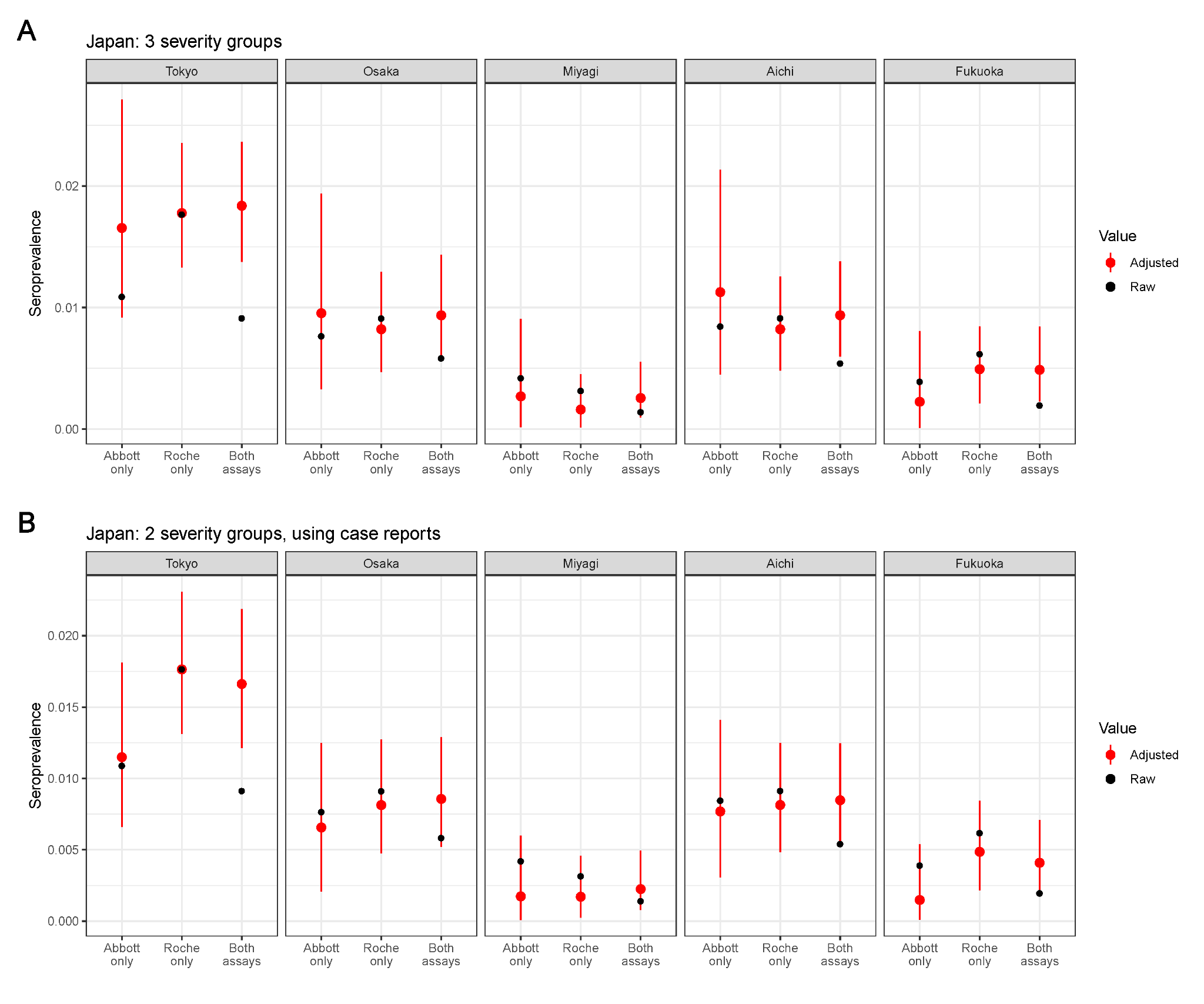

8.	SARS-CoV-2 IgG Architech - Instructions for Use. Available from: <https://www.fda.gov/media/137383/download>

9.	method-sheet-cobas. Available from: <https://www.fda.gov/media/137605/download>

10.	VITROS Immunodiagnostic Products Anti-SARS-CoV-2 IgG Reagent Pack - Instructions for Use. Available from: <https://www.fda.gov/media/137363/download>
